# Stressor-evoked brain activity, cardiovascular reactivity, and subclinical atherosclerosis in midlife adults

**DOI:** 10.1101/2024.02.05.24302236

**Authors:** Javier Rasero, Timothy D. Verstynen, Caitlin M. DuPont, Thomas E. Kraynak, Emma Barinas-Mitchell, Mark R. Scudder, Thomas W. Kamarck, Amy I. Sentis, Regina L. Leckie, Peter J. Gianaros

## Abstract

**Background:** Cardiovascular responses to psychological stressors have been separately associated with preclinical atherosclerosis and hemodynamic brain activity patterns across different studies and cohorts; however, what has not been established is whether cardiovascular stress responses reliably link indicators of stressor-evoked brain activity and preclinical atherosclerosis that have been measured in the same individuals. Accordingly, the present study used cross-validation and predictive modeling to test for the first time whether stressor-evoked systolic blood pressure (SBP) responses statistically mediated the association between concurrently measured brain activity and a vascular marker of preclinical atherosclerosis in the carotid arteries.

**Methods:** 624 midlife adults (aged 28-56 years, 54.97% female) from two different cohorts underwent two information-conflict fMRI tasks, with concurrent SBP measures collected. Carotid artery intima-media thickness (CA-IMT) was measured by ultrasonography. A mediation framework that included harmonization, cross-validation, and penalized principal component regression was then employed, while significant areas in possible direct and indirect effects were identified through bootstrapping. Sensitivity analysis further tested the robustness of findings after accounting for prevailing levels of cardiovascular disease risk and brain imaging data quality control.

**Results:** Task-averaged patterns of hemodynamic brain responses exhibited a generalizable association with CA-IMT, which was mediated by an area-under-the-curve measure of aggregate SBP reactivity. Importantly, this effect held in sensitivity analyses. Implicated brain areas in this mediation included the ventromedial prefrontal cortex, anterior cingulate cortex, insula and amygdala.

**Conclusions:** These novel findings support a link between stressor-evoked brain activity and preclinical atherosclerosis accounted for by individual differences in corresponding levels of stressor-evoked cardiovascular reactivity.

## INTRODUCTION

Acute psychological stressors – defined in a stress typology framework^1,2^ as short-term demands that are appraised as taxing or exceeding one’s coping resources^3^ – typically raise blood pressure (BP) and alter other parameters of cardiovascular physiology in most people^4–7^. There are wide and stable (phenotypic) differences across people, however, in the magnitude, patterning, duration, and direction of stressor-evoked cardiovascular – particularly BP – reactions^8^. According to several conceptual frameworks on psychological stress, BP reactions to acute stressors arise from predictive processes that are instantiated in brain systems for visceral control, which are thought to adjust cardiovascular physiology in order to meet the anticipated metabolic demands that are necessary to cope with acute stressors^9–12^. Notably, some people have a ‘trait-like’ phenotype to exhibit larger than average rises in BP that may exceed the metabolic demands of a stressor^9^. When assessed by standard stressor batteries and testing paradigms, the magnitude of stressor-evoked BP reactions generalizes from the lab to predict those measured in daily life via ambulatory BP monitoring^8,13^. The latter evidence has been interpreted to support the possibility that a phenotype to repeatedly express large-magnitude stressor-evoked BP reactions over the lifecourse may confer cardiovascular risk via their cumulative pathophysiological effects on the vasculature.

In the context of cardiovascular health, for example, epidemiological evidence demonstrates that greater stressor-evoked BP reactivity predicts vascular pathology^14,15^ and early death^16^, consistent with the interpretation that large BP reactions to acute stressors may eventuate in vascular risk – reflected by arterial changes and dysfunction^17–24^. These pathogenic effects may result from repeated pressor influences that injure the endothelial layer of blood vessels via turbulent (non-laminar) blood flow and shear stress^6,25^. At the vascular level, such injury may increase the permeability of the endothelium to lipoproteins, promote the release of mitogenic substances, contribute to the proliferation of intimal smooth muscle cells, and disrupt the lipid metabolism of endothelial cells^26–28^. The effects of a phenotype to exhibit large-magnitude stressor-evoked BP reactivity may thus manifest as endothelial damage and dysfunction, as well as a hypertrophy or thickening of arteries and other blood vessels^6,29^. Consistent with these interpretations, exaggerated stressor-evoked BP reactivity predicts endpoints of the latter pathological changes, including hypertension and a surrogate marker of preclinical vascular disease; namely, carotid artery thickening^16,30,31^.

In parallel to the latter epidemiological findings on preclinical vascular disease, human brain imaging studies have identified functional patterns of neural activity that predict individual differences in stressor-evoked BP reactivity, particularly patterns that have been localized to what have been termed *visceral control circuits*^9^.

Visceral control circuits encompass evolutionarily conserved networks of brain areas spanning the medial prefrontal cortex, anterior cingulate, insula, hippocampus, amygdala, thalamus, hypothalamus, periaqueductal gray, and brainstem cell groups. These circuits control and coordinate *autonomic*, *neuroendocrine*, *hemodynamic*, and *immune* activity across a range of behavioral states that affect cardiovascular physiology and pathophysiology, especially in relation to stressful and emotional experiences that may confer cardiovascular risk. In these regards, visceral control circuits are brain systems that may orchestrate psychological *appraisal* processes of stressors that are calibrated with peripheral physiology to support adaptive action and stressor coping behaviors. Accordingly, visceral control circuits are thought to be capable of centrally orchestrating behavioral influences on cardiovascular risk via peripheral physiological mechanisms, such as acute stressor-evoked BP reactivity^9^.

What has not yet been firmly established, however, is the extent to which stressor-evoked BP reactivity per se statistically explains the possible association of functional activity within visceral control circuits and established subclinical markers of cardiovascular risk, particularly carotid artery thickening. Accordingly, the present study used a predictive machine-learning approach to test the hypothesis that multivariate features of stressor-evoked activity within visceral control circuits of the brain are associated with carotid artery thickening in part via stressor-evoked BP reactivity within a statistical mediation model. Participants from two cohort studies of the neural correlates of stress physiology and cardiovascular risk (total N = 624 midlife adults; aged 28-56 years [54.97% female]) completed a battery of psychological stress tasks designed to measure reliable individual differences (phenotypes) in self-reported and cardiovascular reactivity to acute stressors while global hemodynamic responses in the brain were monitored using fMRI^32–35^. For completeness of reporting, prediction models tested a range of different measures of cardiovascular stress reactivity and physiology.

## MATERIALS AND METHODS

### Participants

Cross-sectional data used herein were from midlife and community-dwelling adults who participated in two different studies with harmonized data collection protocols: The Pittsburgh Imaging Project (PIP), which included 325 individuals (aged 30 – 51 years; 163 women and 162 men; 226 identifying as white; 79 identifying as Black or African American; 15 identifying as Asian or Asian American; three identifying as multiracial; and two reporting other racial identities), and the Neurobiology of Adult Health (NOAH), which included 299 individuals (aged 28 – 56 years; 180 women, 110 men, and one identifying as neither woman nor man; 254 identifying as white; 18 identifying as Black or African American; 19 identifying as Asian or Asian American; seven identifying as multiracial, and one reporting another racial identity). Informed consent was obtained from all participants, and approval was granted by the University of Pittsburgh Human Research Protection Office for PIP (Protocol ID: 07110287) and NOAH (19030012).

Recruitment methods and related methodological details for the PIP cohort have been published^33,36^. Exclusionary criteria for the PIP cohort included: aged < 30 or > 50 years; metallic or other implants unsafe for MRI; for women, pregnancy; any self-reported history of CVD; any self-reported history of a neurological disorder; current treatment for or self-reported diagnoses by a healthcare professional of a psychiatric condition; consuming alcohol equaling or exceeding five servings three or more times/week; regular use of over-the-counter or prescribed medications with autonomic, cardiovascular, or neuroendocrine effects, including daily use of corticosteroid inhalers; any current treatment for hypertension or having a resting BP exceeding 140/90 mmHg; use of any psychotropic medications (e.g., antidepressants); history of metal exposure (e.g., welding); and color blindness.

Recruitment methods for the NOAH cohort involved (a) mass electronic and print mailings to residents of Allegheny County, PA; (b) radio, electronic (e.g., Craig’sList), and print advertisements in public places (e.g., Port Authority Buses, local newspapers, community and park announcement boards); and (c) direct solicitation from the participant registries of the University of Pittsburgh’s Clinical and Translational Science Institute Pitt+Me Registry and University Center for Social and Urban Research Regional Research Registry. Exclusionary criteria for the NOAH cohort included: aged < 28 or > 56 years; self-reported use of medications with central or peripheral autonomic effects on one or more occasions in the 14 days prior to testing (including antihypertensive or cardiac medications, antipsychotic medications, protease inhibitors or other anti-HIV medications, insulin, chemotherapy agents, immunosuppressants, prescription weight-loss medications and ephedrine); regular use (i.e., use on seven or more days in the 14 days prior to testing) of anti-anxiety medications, sleep medications, asthma medications and allergy inhalants, antidepressant medications, glucocorticoids, medical marijuana, more than two non-insulin medications for diabetes; consuming 35 or more alcoholic drinks in the last seven days; consuming six or more alcoholic drinks on three or more occasions in the past seven days; use of illicit drugs on seven or more days in the past two weeks; major medical conditions, including CVD, severe hypertension (SBP/DBP > 160/and/or >100 mmHg); cancer (treatment in last 12 months, except for non-melanoma skin cancer), liver disease, chronic kidney disease, and Type I diabetes; self-reported history of a major neurological disorder or brain injury resulting in ongoing symptoms or cognitive impairment (e.g., multiple sclerosis, cerebral palsy, major head injury); history of schizophrenia or bipolar disorder; lung disease requiring regular or ongoing drug treatment; weight loss surgery within the past five years; for women, pregnancy; regular use of an assistive walking device; non-fluency in English; visual impairments affecting comprehension of printed text or text on a computer screen; color blindness; contraindications for MRI; night shift employment on a frequent basis (operationally defined as working half or more of employment hours in a full workday between midnight and 8 am, and this occurring more than 12 times during the past year); and lack of reliable access to a telephone throughout the day (home, work, or cell phone).

### Cardiovascular Risk Factors

At the time of initial testing, participants in both studies underwent assessments of seated resting blood pressure, waist circumference, and body mass index, as well as fasting glucose and lipid levels. Following guidelines of the American Heart Association, seated resting blood pressures (BPs) were obtained with an oscillometric device (PIP: Critikon Dinamap 8100, Johnson & Johnson, Tampa, FL; NOAH: Omron IntelliSense© BP Monitor, model HEM-907XL, Omron Healthcare Inc). A total of three BPs were taken after an acclimation period, with the average of the last two of the three BPs being used to compute resting systolic (SBP) and diastolic (DBP) blood pressures. Participants’ waist circumference was measured at the level of the umbilicus to the nearest 1/2 centimeter at end expiration. Height was measured by a vertical-mounted stadiometer (with shoes off), and weight was measured in kg.

Similarly, at the time of initial testing, participants underwent fasting phlebotomy for the assessment of glucose, insulin, total cholesterol, triglycerides, and high-density lipoprotein (HDL) cholesterol. If participants were unable to comply with pre-phlebotomy instructions, they were rescheduled. Along with other demographic and anthropometric variables, these measures were used to describe and characterize the study sample and to derive the 10-year ASCVD risk score^37^.

### Carotid Artery Intima-Media Thickness by Ultrasonography

Participants in both cohorts underwent the same carotid artery ultrasonography protocol, which was performed by a registered vascular technologist in the laboratory of co-author E. B-M. Specifically, each participant laid supine with the head tilted at 45°, and, using an Acuson Antares scanner (Acuson-Siemens, Malvern, PA), the technologist performed scout views of the left and right carotid arteries in both the transverse and longitudinal planes. A region-of-interest encompassing the artery walls was identified for more focused B-Mode imaging of three carotid areas: (1) the near and far walls of the distal common carotid artery (1 cm proximal to the carotid bulb, measured in duplicate on each side and averaged across images and sides); (2) the far wall of the carotid bulb (defined as the point where the near and far walls of the common carotid are no longer parallel and extending to the flow divider); and (3) the first cm of the internal carotid (defined distally from the edge of the flow divider). For the three carotid areas (common, bulb, and internal), an optimal image was digitized for later scoring with automated edge detection software (Artery Measurement System; Goteborg University, Gothenburg, Sweden). The software was used to draw two lines: one along the lumen-intima interface and one along the media-adventitia interface. The distances between the line-identified interfaces were measured in 1 cm segments, generating one measurement (in mm) for each pixel in each segment (approximately 140 measurements total). As pre-registered, the primary dependent (Y) variable in the present analyses corresponded to the mean average carotid artery intima media thickness averaged over the far walls of the carotid bulb and common carotid artery, henceforth CA-IMT.

### Cardiovascular reactivity

During fMRI testing, BP was measured using MRI compatible devices (PIP: Medrad Multigas 9500, Warrendale, PA/Leverkusen, Germany; NOAH: Tesla M3 MRI Patient Monitor by the Mammendorf Institute for Physiology and Medicine, Mammendorf, Germany). The average incongruent condition—minus—average baseline BP = difference was used to compute reactivity. Task-averaged Systolic BP reactivity values, or ΔSBP, were used as the primary mediator variable (M) in our study. Additional analyses that were pre-registered also considered other reactivity metrics, including (a) the area under the curve with respect to ground, or SBP_AUC_g_; and (b) the area under the curve with respect to increase^38–40^, or SBP_AUC_i_, conditional on these alternative metrics of reactivity exhibiting statistical associations with carotid-artery intima-media thickness. Unlike ΔSBP, both area-under-the-curve measures incorporate SBP changes throughout the entire task. As a result, they are capable of capturing more information, such as subject habituation to the task and the total or aggregate load of BP throughout the entire task as opposed to the average peak increase. Moreover, AUC measures have been shown to exhibit better phenotypic test-retest reliability compared to measures like ΔSBP^41^.

### fMRI stressor task battery

The tasks of this battery include a Stroop task and a multi-source interference task (MSIT), each 9 min 20 sec. Both entail processing conflictual information, receiving negative feedback, and making time-pressured responses to unpredictable and uncontrollable stimuli that elicit subjective distress^35^. Briefly, subjects complete four, 52-60 sec blocks of trials in both tasks that define a congruent condition, which are interleaved with four, 52-60 sec blocks of trials defining an incongruent condition. Conditions are preceded by a 10-17 sec fixation period. In the Stroop, subjects identify the color of target words in the center of a screen by selecting one of four identifier words. Selections are made by pressing one of four buttons on a glove, with each button matching an identifier word on the screen (e.g., thumb button 1 = identifier word on the left, etc.). During congruent Stroop trials: (1) targets are in colors congruent with the target words, and (2) identifiers are in the same colors as targets. During incongruent Stroop trials: (1) targets are in colors incongruent with the targets, and (2) identifiers are in colors incongruent with the colors that the identifiers name. In the MSIT, subjects in the NOAH study selected a number that differed from four others by pressing one of four buttons on the glove, with each button matching a number on the screen (thumb button 1=number 1, etc.). During congruent MSIT trials, targets were in a position compatible with their position on the glove. During incongruent MSIT trials, targets were in a position incompatible with their glove position. In the PIP study, subjects were presented with three number stimuli and not four as in NOAH. One methodological difference is that while the PIP study utilized zeros in the MSIT to indicate an incompatible position in the congruent condition (e.g. [1 0 0]), NOAH used one number between 1 and 4 (e.g. [1 2 2 2]) during this condition. In incongruent conditions of both tasks in both cohorts, accuracy was held to ∼50% by adjusting inter-trial intervals (ITIs). Thus, consecutive accurate performance in an incongruent condition prompted shorter ITIs. Conversely, less accurate performance lengthened ITIs. To control for motor response differences between conditions, the number of trials in the congruent condition was yoked to the number completed in the incongruent condition. To implement yoking, (1) an incongruent block is administered first, and (2) congruent condition trials appear at the mean ITI of the preceding incongruent block.

### MRI Data Acquisition and Preprocessing

Imaging in the PIP cohort was conducted using a 3-Tesla Trio TIM scanner (Siemens, Erlangen, Germany). Functional BOLD data for the Stroop and MSIT tasks were acquired with a gradient echo-planar imaging sequence by these parameters: time to repetition/time to echo=2000/28 ms; matrix resolution=64 x 64; field of view=205 x 250 mm; and flip angle=90°. Each volume was 3 mm in thickness, with no gap (280 task volumes in total, excluding 4 discarded volumes). Prior to functional imaging, a T1-weighted magnetization prepared rapid gradient echo (MPRAGE) structural image was obtained by these parameters: repetition time = 2100 msec; inversion time = 1100 msec; echo time = 3.31 msec; flip angle = 8°. There were 192 sagittal slices (1 mm thick, no spaces between slices) having a matrix size = 256 x 208 pixels (field-of-view [FOV] = 256 x 208 mm).

In the NOAH study, imaging was conducted using a 3-Tesla PRISMA scanner (Siemens, Erlangen, Germany), which was equipped with a 64-channel head coil. Prior to functional imaging, a T1-weighted magnetization prepared rapid gradient echo (MPRAGE) structural image was obtained by these parameters: repetition time = 2300 msec; inversion time = 900 msec; echo time = 1.99 msec; flip angle = 9°. There were 176 sagittal slices (1 mm thick, no spaces between slices) having a matrix size = 176 x 176 voxels (field-of-view [FOV] = 256 x 256 mm). Functional BOLD image acquisition parameters for the Stroop and MSIT tasks were: matrix size = 106 × 106 voxels (FOV = 212 × 212 mm), TR = 2000 ms, TE = 30 ms, and FA = 79°. 69 slices per volume were collected along an anterior-to-posterior encoding direction. Each volume was 2 mm in thickness, with no gap (280 task volumes in total, excluding 4 discarded volumes).

Individual fMRI data for both cohorts and tasks underwent the same preprocessing pipeline using statistical parametric mapping software (SPM12; http://www.fil.ion.ucl.ac.uk/spm). For spatial preprocessing, T1-weighted MPRAGE images were classified into six tissue types. Biased-corrected and deformation field maps were then computed. Functional images were realigned to the first image of the series by a 6-parameter rigid-body transformation, using the re-slice step to match the first image on a voxel-by-voxel basis. Before realignment, slice-timing correction was applied to account for acquisition time variation. Realigned images were co-registered to each participant’s skullstripped and biased-corrected MPRAGE image. Co-registered images were normalized to Montreal Neurological Institute (MNI) space. In within-individual fMRI analyses, univariate general linear models (GLMs) were estimated to compute contrast maps used for prediction and mediation analysis described below in the analysis plan. Task blocks were modeled by rectangular waveforms convolved with the default hemodynamic response function in SPM12. These regressors modeled blocks (i.e. fixation, incongruent condition, congruent condition). In each GLM, the six realignment parameters from pre-processing were included as nuisance regressors and low-frequency artifacts were removed by a high-pass filter (187 s). Error variance was estimated and then weighted by restricted maximum likelihood estimation, as implemented in the RobustWLS toolbox, v4.0. Subsequently, each estimated task condition parameter was smoothed by a 6mm full-width-at-half-maximum (FWHM) Gaussian kernel. Linear contrasts, computed as *‘Incongruent vs. Congruent’* condition effects and averaged across Stroop tasks and MSIT, constituted the independent variable (X) in our main analysis (see next section).

Exclusion criteria of individual fMRI data included lack of spatial (brain) coverage, incomplete sequence acquisition, experimental error, equipment malfunction during participant testing, and lack of participant task comprehension after reviewing task performance and experimenter notes.

### Mediation analysis

A mediation framework was used to test the association between the task-averaged whole-brain fMRI ‘Incongruent vs. Congruent’ activity maps (the multivariate independent variable, X) and carotid artery intima-media thickness (the univariate outcome variable, Y), mediated by task-averaged in-scanner SBP reactivity (the univariate mediator, M). In order to accomplish this and following Baron and Kenny’s steps^42^, the X-to-Y (“Total Effect”) or *c* path was first assessed, i.e. the relation between the task-averaged stressor-evoked fMRI activation patterns and the carotid artery intima-media thickness. Next, the X-to-M or *a* path was estimated, i.e. the effect of the task-averaged stressor-evoked fMRI activation patterns on task-averaged stressor-evoked SBP reactivity. Finally, the *b* and *c’* paths were estimated, which are defined as the amount of variability of Y that M and X respectively explain when taken together in the same model ([X+M]-to-Y). As a result, statistical evidence for a *c’* path indicates that X has a direct effect on Y. Similarly, statistical evidence for both *a* and *b* paths indicates that X has an effect on Y mediated by M (“Indirect effect”). To note, the whole mediation scenario was conducted separately for each potential mediator (ΔSBP, SBP_AUC_g_ and SBP_AUC_i_).

In the current modeling framework, the aforementioned mediation scenario was tested using a machine-learning approach. Specifically, the reliability of each effect was assessed using a nested k-fold cross-validation, consisting of (a) an ‘inner loop’ 5-fold cross-validation to optimize the predictive models, and (b) an ‘outer loop’ 10-fold cross-validation to determine the predictive generalizability of these models. This data splitting took place separately in each mediation analysis conducted and in a stratified fashion, so M and Y variables did not differ statistically between training and test sets. Stratification also took cohort information into account, ensuring that both studies were included during training and testing while maintaining the original proportion between them. The whole cross-validation procedure was also repeated with five different seeds to account for the variability when splitting the data.

With regards to the predictive model, because each mediation step entails a high degree of dimensionality (number of features greater than the number of observations), a penalized principal component regression was employed. Such an estimator involved the following sequential steps:

1. Dimensionality reduction by PCA to the input fMRI data X:

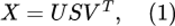 where *Z* = *US* corresponds to the projected values of *X* into the principal component space and *V^T^* an orthogonal matrix that contains the principal axes in feature space.
2. Regression of the response variable (Y or M, depending on the model) onto Z, with the estimation of the *β* coefficients given by the following cost function

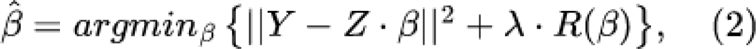 where R(beta) is a penalty term weighted by a parameter lambda.
3. Projection of the estimated *β* coefficients (in the principal component space) back to the original feature space to generate a weight pattern *ŵ* as follows

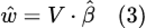
4. Use of this weight pattern to yield a final prediction:

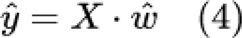

In addition, both L1 (also known as Lasso, R(beta) = ||*β*||) and L2 (also known as Ridge, R(beta) = ||*β*||^2) regularization forms were tested. The inner 5-fold cross-validation loop, applied **only** to the training set, was utilized to find the optimal value of λ out of a sequence of 1000 different values. Specifically, the performance of each λ was evaluated by calculating the mean squared error (MSE) between predicted and observed outcome variables in respective validation samples. The optimal value of λ corresponded to that with the lowest MSE, which was also used to deploy an optimized predictive model via refitting the entire sample of the inner loop with such λ parameter. The above dimensionality reduction step was applied only to X, i.e. the principal component space was exclusively based on the whole-brain fMRI activation data. This also ensured that X and M information were not mixed together in the last model tested ([X+M]-to-Y).

The generalizability of each optimal predictive model (i.e. X-to-Y, X-to-M and [X+M]-to-Y models) was evaluated by comparing predicted with observed outcome variables in the test samples. Here, the similarity between predicted and observed values was summarized by Pearson correlation coefficients and corresponding 95% bootstrapped confidence intervals (CI) and p-values. Following guidelines for predictive modeling^43^, variance in observed values explained by predicted values (R^2^) was calculated by the sums-of-squares formulation. Furthermore, to estimate the generalizability of the mediation effect the following measure was adopted: 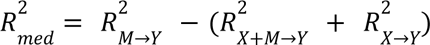. This is based on the effect sizes of each individual model in the mediation analysis framework^44^ and therefore, easily evaluated out-of-sample. Finally, the evidence for each effect was quantified using Bayes factors, BF_01_, which reflect the ratio between the probability of the alternative (existence of a positive correlation between predicted and observed values) and the null hypotheses (absence of or negative correlation between predicted and observed values).

Individual features that reliably contributed to both direct effects (or *c’* path) and indirect effects (i.e. the products *a·b*) were determined by bootstrap resampling (5000 resamples), with corrections for a false discovery rate of 0.05^45^. This analysis was performed on the principal component space to increase statistical power. However, the *β* coefficients estimated directly from a bootstrapped sample X* would not be calculated in the same principal component space as that of the original data X. In order to solve this, let and 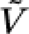 be respectively 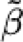 the regression coefficients for the principal components and their transformation matrix to the original feature space, both estimated from the bootstrapped sample. Following eq. 3, a weight pattern *w\** in the voxel space was first obtained. This weight pattern was then transformed to *β*\* using the original orthogonal matrix V, thus ensuring that the regression coefficients from a bootstrapped sample were estimated in the same principal component axes computed from X. The resulting subsets of significant *c’* and *a·b* coefficients from this bootstrapping procedure were then projected back to the voxel space to yield whole-brain patterns of direct and indirect effects. Finally, in order to ensure that these fMRI activity patterns clearly indicated a direct relation with the response variable, a transformation to encoding weight maps was performed^46^.

Importantly, because of the use of two different cohorts, harmonization techniques were applied with the aim of reducing the differences between NOAH and PIP. In particular, for neuroimaging data (X), ComBat was adopted, which is a Bayesian-based statistical method that has been shown to adjust for location and scale effects from different data sources^47^. M and Y were similarly harmonized by standardizing them within cohorts.

Finally, it is important to note that each of the different steps that led to the final predictions (harmonization, dimensionality reduction, etc.) were all part of the analytical pipeline and more importantly, were embedded within the cross-validation procedure, thus preventing any data leakage.

### Ancillary analysis

In order to test whether observed results could be explained by prevailing levels of overall cardiovascular disease risk across participants, 10-year Atherosclerotic Cardiovascular Disease (ASCVD) risk scores were used as control variables to examine the influence on predicted vs. observed correlation values. This analysis was contingent on the existence of a significant correlation between ASCVD risk scores and carotid artery intima media thickness, and for any of the cardiovascular reactivity measures tested where such a significant correlation also existed. Additionally, sensitivity analyses assessed the influence of functional brain imaging data quality by excluding subjects with excessive head motion during any of the task acquisitions (average framewise displacement greater than 0.5 mm using the formula in ^48^).

### Preregistration and Availability of Code and Data

Hypotheses and planned analyses were pre-registered at Open Science Framework (OSF) on 01 June 2022 (https://osf.io/j278q). Subsequently created data files and analysis scripts are available at GitHub (https://github.com/CoAxLab/sbp-imt-mediation). To the authors’ awareness, this is the first study to investigate the cross-sectional association between stressor-evoked brain activity patterns and CA-IMT mediated by changes in BP.

## RESULTS

### Sample characteristics

Descriptive information and cohort comparisons of means (t-tests) and variances (Bartlett tests) for demographic and cardiovascular measures are summarized in Table 1. In particular, carotid artery intima-media thickness (CA-IMT) and cardiovascular reactivity measures (ΔSBP; SBP_AUC_g_; SBP_AUC_i_) differed statistically between cohorts. These latter statistical differences were likely attributable to inter-device variability for BP monitoring during fMRI. Accordingly, these variables were harmonized in the predictive analysis by being standardized within cohorts. This step was embedded within the cross-validation procedure to avoid data leakage.

**Table 1.** Data characteristics and descriptive statistics for the continuous variables in the study. Statistical differences in location (mean) and scale (variance) between cohorts were assessed using a two-sample t-test and Bartlett’s test respectively. In bold, the significant (α=0.05) false discovery rate corrected p-values (q-values).

|  | PIP<br>(N=325, 163 women) |  | NOAH<br>(N=299, 180 women) |  |  |  |
| --- | --- | --- | --- | --- | --- | --- |
|  | Mean<br>[95 % CI] | Standard<br>Deviation | Mean<br>[95 % CI] | Standard<br>Deviation | q-value<br>(location) | q-value<br>(scale) |
| Age (years) | 40.89<br>[40.21-41.60] | 6.24 | 42.31<br>[41.24-43.6] | 8.64 | **0.03** | **<0.01** |
| Number of school years completed | 16.67<br>[16.33 - 17.04] | 3.31 | 17.63<br>[17.32-17.96] | 2.85 | **<0.01** | **0.01** |
| Weight (lb) | 174.13<br>[170.31-177.98] | 35.44 | 168.03<br>[164.26-172.29] | 37.99 | 0.05 | 0.25 |
| Waist (Inches) | 38.04<br>[34.95 - 43.68] | 47.66 | 36.81<br>[36.16 - 37.43] | 5.56 | 0.64 | **<0.01** |
| Height (Inches) | 67.46<br>[67.04 - 67.88] | 3.71 | 66.84<br>[66.42 - 67.24] | 3.66 | 0.05 | 0.81 |
| Glucose (mg/dL) | 88.15<br>[86.91 - 89.36] | 11.58 | 87.33<br>[86.40 - 88.34] | 8.51 | 0.36 | **<0.01** |
| Triglyceride (mg/dL) | 94.05<br>[87.68 - 100.75] | 57.46 | 97.88<br>[88.88 - 110.34] | 89.89 | 0.57 | **<0.01** |
| Hdl (mg/dL) | 50.89<br>[49.16 - 52.72] | 16.17 | 57.74<br>[56.05 - 59.53] | 14.88 | **<0.01** | 0.2 |
| Insulin (μU/mL) | 8.92<br>[8.06 - 9.81] | 6.50 | 6.72<br>[6.04 - 7.38] | 6.01 | **<0.01** | 0.25 |
| BMI (kg/m<sup>2</sup>) | 26.88<br>[26.38 - 27.42] | 5.08 | 26.27<br>[25.59 - 26.87] | 5.35 | 0.17 | 0.38 |
| CA-IMT (mm) | 0.64<br>[0.63 - 0.65] | 0.10 | 0.60<br>[0.59 - 0.62] | 0.13 | **<0.01** | **<0.01** |
| Clinic-SBP (mm Hg) | 121.01<br>[119.76 - 122.13] | 10.66 | 114.49<br>[112.85 - 116.05] | 13.49 | **<0.01** | **<0.01** |
| ΔSBP (mm Hg) | 4.22<br>[3.57 - 4.83] | 5.75 | 2.84<br>[2.04 - 3.68] | 6.78 | **0.02** | **0.01** |
| SBP_AUC<sub>g</sub><br>(mm Hg · s) | 1014.78<br>[1003.33 - 1025.46] | 100.20 | 943.60<br>[929.78 - 958.16] | 118.43 | **<0.01** | **0.01** |
| SBP_AUC<sub>i</sub><br>(mm Hg · s) | 27.85<br>[23.72 - 32.45] | 39.13 | 17.35<br>[11.60- 23.38] | 47.41 | **0.01** | **<0.01** |
| ascvd_10y | 1.90<br>[1.74- 2.11] | 1.69 | 1.55<br>[1.42 - 1.69] | 1.20 | **0.01** | **<0.01** |

Next, we assessed the associations between CA-IMT, SBP reactivity metrics, and CVD risk factors. As shown in Figure 1, greater CVD risk (e.g., as per ASCVD scores) correlated positively with CA-IMT (PIP: r(300)=0.32, q<0.001; NOAH: r(283)=0.388, q<0.001), as well as positively with SBP_AUC_g_ (PIP: r(296)=0.286, q<0.001; NOAH: r(251)=0.251, q<0.001). Similar findings were observed for several other CVD risk factors; however, these associations did reliably replicate across the PIP and NOAH cohorts. Notably, SBP reactivity as indexed by ΔSBP did not appear to reliably correlate with CVD risk factors (PIP: r(297)= −0.01, q=0.928; NOAH: r(245)=0.047, q=0.592). A similar situation was observed for SBP_AUC_i_ (PIP: r(296)=-0.012, q=0.927; NOAH: r(251)=0.093, q=0.227). As a result, and following the pre-registration plan, only comparisons of predicted vs observed values using SBP_AUC_g_ as a mediator between stressor-evoked fMRI activity and CA-IMT were tested, controlling for prevailing levels of CVD risk indexed by ASCVD. For completeness, pairwise correlations between all these variables using the combined sample (PIP + NOAH) can be found in Suppl. Figure 1, with similar conclusions reached as per study.

**Figure 1.**
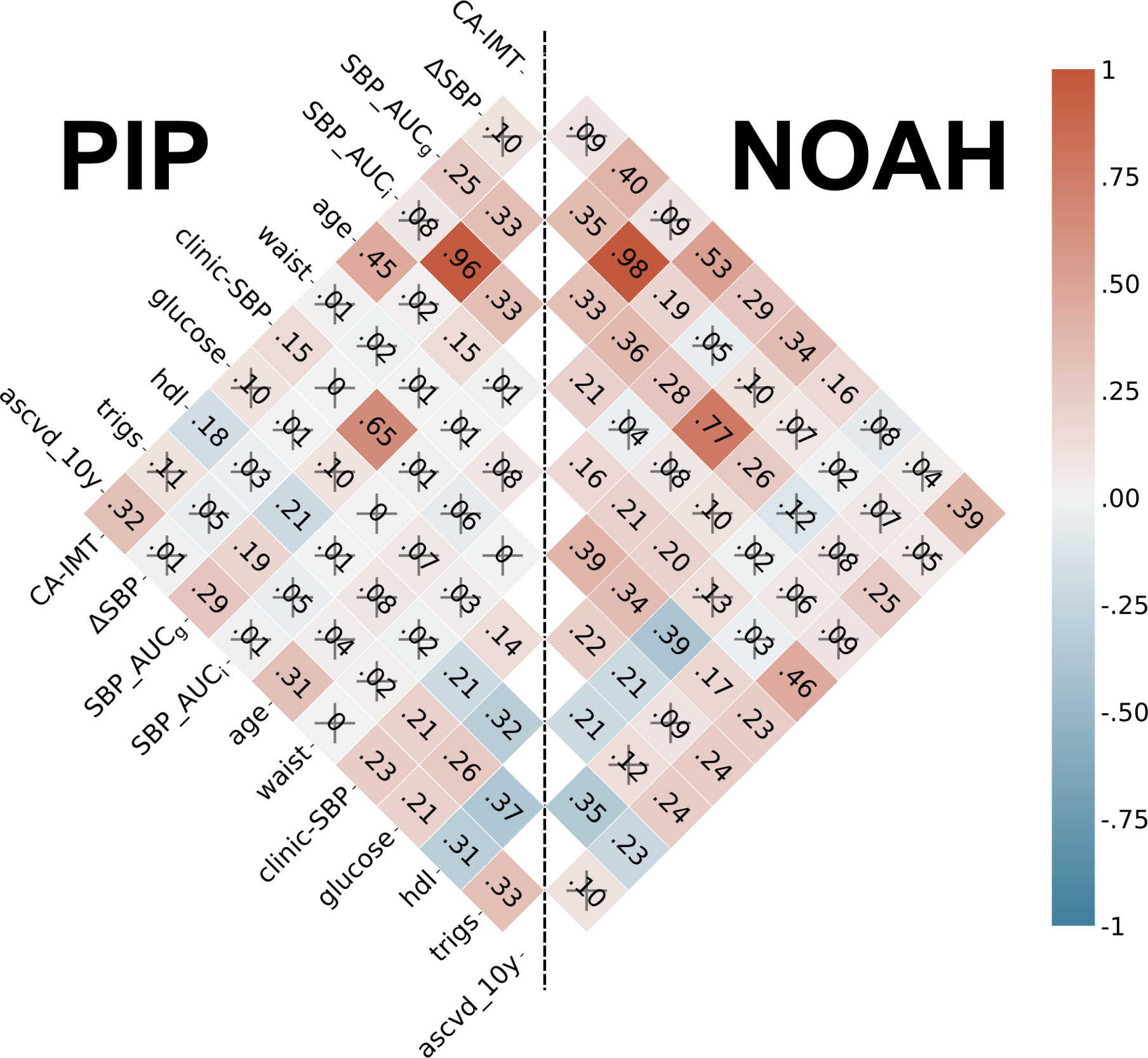
Correlogram. A correlogram involving the main variables in our mediation framework and several cardiovascular disease risk factors. Colors and annotations display the strength of their associations as measured by Pearson’s correlation coefficients. A cross represents those associations whose p-values after false discovery rate correction were above the significance level α=0.05. SBP, systolic blood pressure; HDL, high-density lipoproteins; Trigs, triglycerides; CA-IMT, carotid artery intima-media thickness; mm, millimeter, SBP_AUC_g_, Area under the curve with respect to the ground; SBP_AUC_i_, Area under the curve with respect to increase; ascvd_10y, 10-year atherosclerosis cardiovascular disease risk score.

### Main effects of the stressor-evoked fMRI tasks

Stress-evoked activation patterns were calculated as contrasts in average brain activity between incongruent and congruent trial conditions in both Stroop task and MSIT, and for each cohort (PIP and NOAH). Areas typically implicated in conflict processing were found to be engaged (see Figure 2). This mainly involved positive activation of the dorsomedial prefrontal cortex, anterior cingulate cortex, anterior insula, parietal cortex, basal ganglia, thalamus, and cerebellum; and the negative activation of areas included in the default-mode network, encompassing the ventromedial prefrontal cortex, perigenual anterior cingulate cortex, posterior cingulate cortex, and precuneus. Overall, there was a large voxel-wise spatial similarity in activation patterns (Pearson’s correlation *r* > 0.8) between tasks in PIP, and between cohorts for Stroop task. Spatial similarity rates weakened when the activation pattern of MSIT in NOAH was involved, as a consequence of an overall decrease in observed effect sizes that particularly affected the lack of presence of significantly deactivated areas, i.e. in the default-mode network (e.g. the medial prefrontal and posterior cingulate cortex; Suppl. Figure 2). This finding can be further understood by comparing the participants’ behavior during MSIT across both cohorts. For example, a comparison can be made for the accuracy in correct responses of each participant based on their choices during the tasks (a word selection in Stroop, a number selection in MSIT). Given that they serve as a control condition, high average performance should be observed across the congruent trials relative to the more challenging incongruent trials. However, while PIP participants did show the expected high accuracy (percentage of correct responses) during the congruent (91.08 ± 7.41%) compared to the incongruent (56.05 ± 9.09%) condition, this difference was considerably smaller for NOAH participants (congruent: 64.25 ± 11.38%; incongruent: 53.92 ± 2.51%) as attributable to the difference in the task structure/design. Therefore, relative to PIP participants, those in the NOAH cohort performed approximately 27% less accurately on average during congruent trials, resulting in a similar decreased BOLD activity of the default-mode network relative to incongruent trials and in contrast to what occurs in the PIP cohort. Combat harmonization of the task-averaged activation patterns across both cohorts reduced (but did not entirely eliminate) these spatial differences (See Suppl. Figure 3).

**Figure 2.**
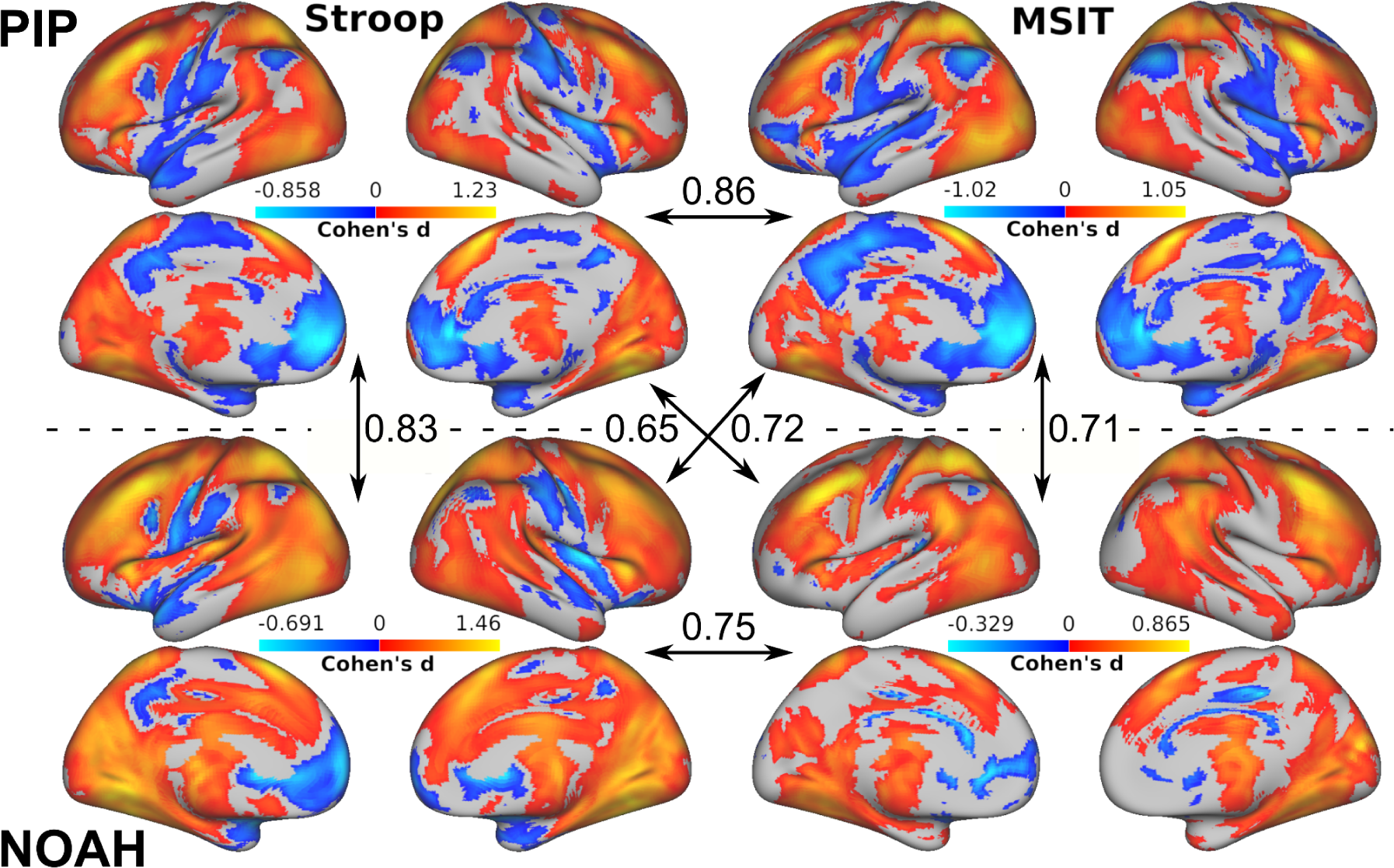
Main effects. For each cohort (PIP and NOAH) the stressor-evoked brain activation patterns at the group level and in terms of the Cohen’s d effect size measure in both fMRI tasks (Stroop and MSIT). Only effects whose p-value is below 0.05 after false discovery rate correction and with a cluster size threshold of *k* > 50 voxels are displayed. Orange colors represent greater average activity in the incongruent condition compared to the congruent condition, and blue colors the opposite. In arrows, the spatial similarity between activation patterns as measured by Pearson’s correlations.

### Prediction of CA-IMT from stressor-evoked brain activation patterns and mediated by cardiovascular reactivity

A mediation analysis employing L2-penalized (i.e., ridge regression) principal component regressions tested the association between stressor-evoked brain activity (input variable; X) and CA-IMT (outcome variable; Y) mediated by several cardiovascular reactivity measures: ΔSBP, SBP_AUC_g_ and SBP_AUC_i_ (mediator variable; M). Figure 3 and Table 2 show that a significant association, in the hold out test sets, as indicated by the average coefficient of determination R^2^ and 95% confidence intervals, was found in the X-to-Y path for the three mediators, as well as in the X-to-M path and the [X+M]-to-Y path. Prediction performances summarized by Pearson’s correlation coefficients can be found in Suppl. Table 1. In addition, predicted vs. observed values across the different cross-validation repeats scatter plots are displayed in Suppl. Figures 4, 5, and 6. As noted in the Methods, the stratification in M and Y within each mediation analysis led to different data partitions and therefore to slightly different prediction rates in the X-to-Y path, even though this model did not involve the mediator. Nonetheless, they all remained within the same confidence intervals.

**Figure 3.**
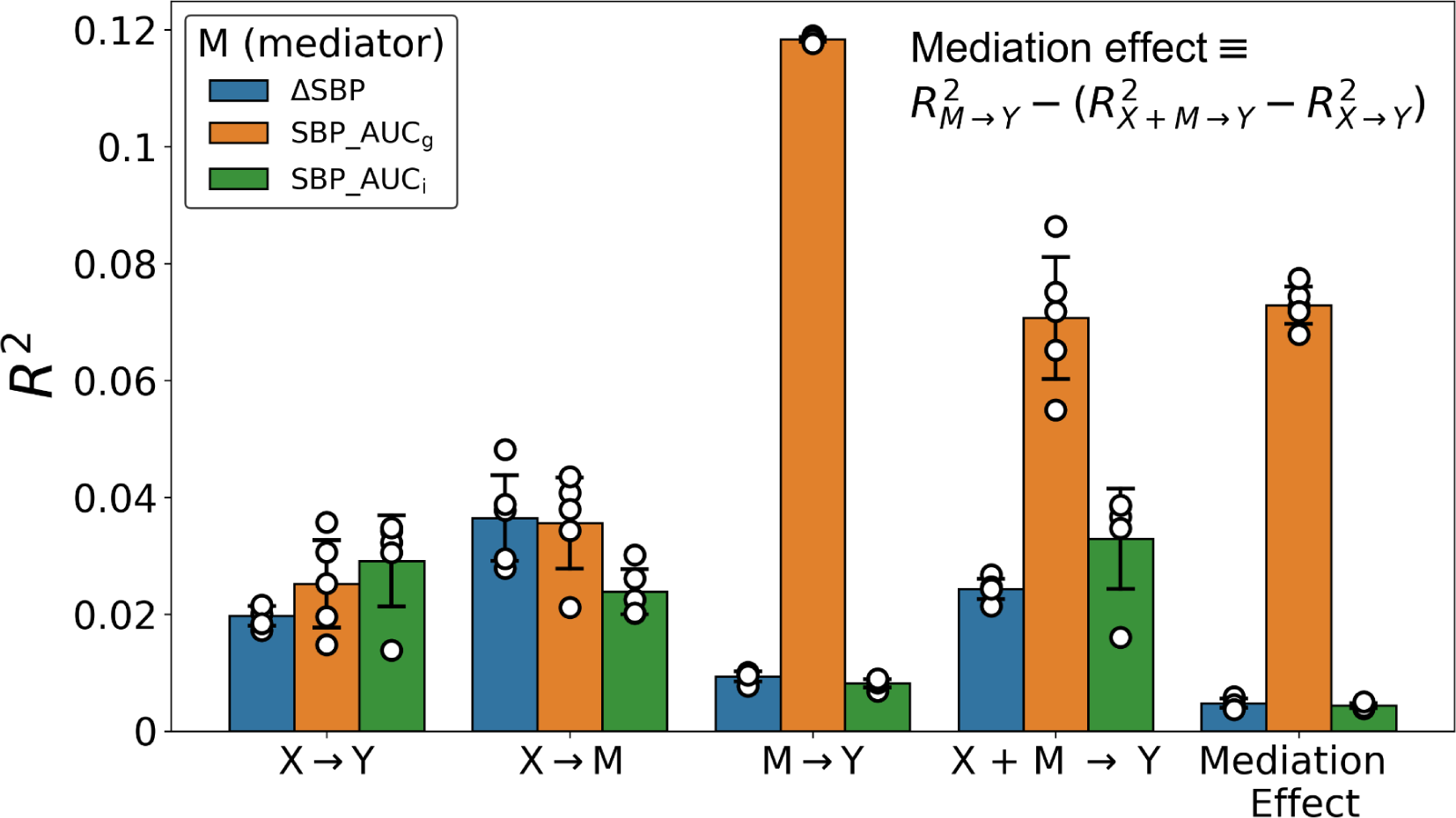
Out-of-sample performances. For each possible mediator variable (M), the out-of-sample performance using a L2-penalized principal component regression and applied to the different paths in the mediation analysis framework. Here, the input variable (X) is voxelwise responses and the output variable (Y) is CA-IMT. Each dot represents the coefficient of determination calculated from the observed .vs predicted values generated from a particular run of the nested cross-validation procedure. In addition, bars and error bars display the average and the 95% Confidence intervals across these values.

**Table 2.** Coefficients of Determination, R^2^. Average Coefficient of Determination, R^2^, and 95% CI for each mediator variable (columns) and model path (rows). The mediation effect was calculated using the R^2^ values in each path as explained in Methods.

| | ΔSBP | SBP_AUC<sub>q</sub> | SBP_AUC<sub>i</sub> |
| --- | --- | --- | --- |
| X-to-Y path | 0.02<br>[0.018 - 0.021] | 0.025<br>[0.018 - 0.033] | 0.029<br>[0.021 - 0.037] |
| X-to-M path | 0.036<br>[0.029 - 0.044] | 0.036<br>[0.028 - 0.043] | 0.024<br>[0.02 - 0.028] |
| M-to-Y path | 0.009<br>[0.009 - 0.01] | 0.118<br>[0.118 - 0.119] | 0.008<br>[0.007 - 0.009] |
| [X+M]-to-Y path | 0.024<br>[0.023 - 0.026] | 0.071<br>[0.06 - 0.081] | 0.033<br>[0.025 - 0.041] |
| Mediation effect | 0.005<br>[0.004 - 0.006] | 0.073<br>[0.070 - 0.076] | 0.004<br>[0.004 - 0.005] |

The previous results showed a generalizable effect for each model that comprises our mediation analysis framework. However, as shown in Figure 3 and Table 2, only SBP_AUC_g_ had a sizeable and reliable mediation effect (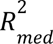 = 0.073, 95% CI [0.070, 0.076]), whereas it was substantially smaller for ΔSBP (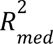 = 0.005, 95% CI [0.005, 0.006]) and SBP_AUC_i_ (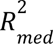 = 0.004, 95% CI [0.004, 0.005]). This can be easily understood by the small variability explained by the latter two mediators in the M-to-Y path (ΔSBP: 0.009, 95% CI [0.009, 0.01]; SBP_AUC_i_: 0.008, 95% CI [0.007, 0.009]; see Figure 3 and Table 2). As a consequence, further analyses concentrated exclusively on SBP_AUC_g_ in testing mediation effects between stressor-evoked brain activation and CA-IMT.

For completeness, we repeated the same analysis using L1-penalized principal component regressions and found similar results, although with an overall decrease in effect sizes (see Suppl. Figures 7, 8, and 9). Thus, findings appear to not depend on the type of penalty applied in modeling.

After conducting bootstrapping (5000 resamples) and correcting for multiple testing using a false discovery rate of 0.05, we identified ten significant principal components for the indirect effects (i.e. *a·b* products), and no significant principal components for the direct effects (*c’* coefficients). In the mediation analysis context, this suggests that the association between stressor-evoked brain activation and CA-IMT appears to be **fully** mediated by SBP_AUC_g_. When we transformed these ten significant components back to the voxel space, we found a (mediated) positive association in areas involving particularly the insula, thalamus, ventromedial prefrontal cortex, anterior cingulate cortex, superior parietal lobe, and vermis (see encoding weight maps in Figure 4). Since the *b* coefficient was always positive, this means that SBP_AUC_g_ increases as incongruency-related brain responses in these areas increases. Conversely, we observed a (mediated) negative association in the dorsomedial prefrontal cortex, angular gyrus, amygdala, cerebellum, and brain stem. Therefore, when incongruency-related brain patterns increase in these areas, SBP_AUC_g_ tends to decrease. Interestingly, there appeared to be a lateralization of the amygdala, since positive associations take place in the left hemisphere and negative associations in the right hemisphere.

**Figure 4.**
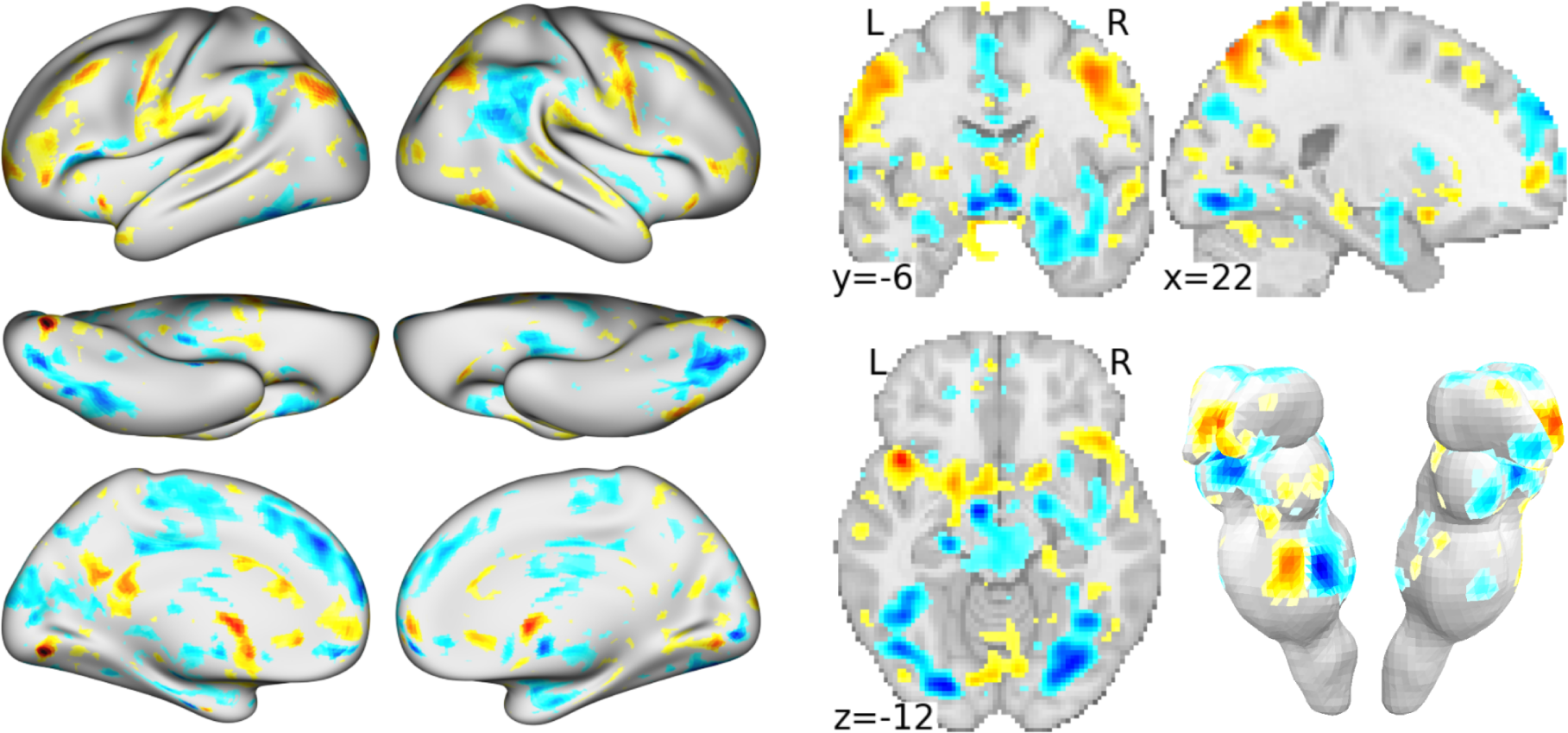
Encoding weight maps of indirect effects. For the scenario with SBP_AUC_g_ as mediator, the encoding weight maps obtained from transforming back to voxel space those principal components whose *a·b* products were significant based on bootstrapping (5000 resamples) and after correcting for a false discovery rate of 0.05. Warm and cool colors represent a positive and negative mediated association between stressor-evoked brain activity and CA-IMT respectively. For visualization purposes, only weights with |z| > 1 after spatial standardization are displayed.

Finally, Suppl. Figure 10 displays the brain patterns of encoding weights from the X-to-M path for each mediator variable. We include them here for comparison, retaining all their principal components information. As expected, due to the high correlation between ΔSBP and SBP_AUC_i_ (see Figure 1 for the correlation values in both cohorts), their weight maps in the X-to-M path also exhibit a large spatial similarity (r=0.96). In contrast, their spatial similarity with the weight map for SBP_AUC_g_ substantially decreased (ΔSBP: r=0.42, SBP_AUC_i_: r=0.43). These differences appear to be triggered by an overall presence of negative associations in the dorsomedial prefrontal cortex, anterior cingulate cortex and angular gyrus for SBP_AUC_g_, unlike ΔSBP and SBP_AUC_i_ where these associations are positive.

### Ancillary testing

An overall decrease in explained variability was observed after including the 10-year ASCVD risk score as a control variable in the scenario involving SBP_AUC_g_ as a mediator. Nevertheless, such associations remained significant for the X-to-M path (R^2^ = 0.022, 95% CI [0.014, 0.022]) and the [X+M]-to-Y path (R^2^ = 0.039, 95% CI [0.027, 0.05]). Only for the X-to-Y path two repetitions of the cross-validation procedure no longer yielded a significant correlation in observed vs predicted values. Altogether, a significant mediation effect was still observed (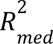 = 0.046, 95% CI [0.042, 0.049]). Therefore, we can conclude that the prevailing values of cardiovascular disease risk factors do not drive this mediation effect between stressor-evoked brain activity and CA-IMT.

Finally, we performed a sensitivity analysis by concentrating exclusively on subjects with low in-scanner head motion, defined as having an average framewise displacement lower than 0.5 mm in both fMRI tasks. This resulted in the exclusion of 27 subjects (23 belonging to PIP, and 4 to NOAH). A mild improvement in performance rates was overall observed (see Suppl. Figure 11), although conclusions previously reached remained the same.

## DISCUSSION

In this study we tested whether whole-brain hemodynamic activity patterns, evoked by two aversive information conflict tasks (Stroop and MSIT), were associated with CA-IMT, a vascular marker of preclinical atherosclerosis. We also tested whether this association was statistically mediated by three different measures of cardiovascular reactivity. In order to accomplish this, we used a multi-cohort dataset comprising ∼600 subjects and the combination of harmonization techniques and penalized principal component regressions to estimate generalizable out-of-sample predictions from a repeated nested cross-validation procedure. Within a mediation analysis framework, we found that stressor-evoked brain activity patterns explained ∼2% of total variability of CA-IMT (the X-to-Y path). The same activation patterns were able to predict cardiovascular reactivity (the X-to-M path), with a variability between 4% and 2% depending on the mediator variable. In addition, when brain activation patterns and cardiovascular reactivity were considered together (the [X+M]-to-Y path), prediction rates of CA-IMT increased, reaching the maximum explained variability (around 8%) for the case of SBP_AUC_g_ as a mediator. Collectively, the present findings suggest that the association between stressor-evoked brain activity and CA-IMT is likely to be mediated most reliably by SBP_AUC_g_, and mainly via the activation of areas such as the insula, thalamus, ventromedial prefrontal cortex, superior parietal lobe; and the deactivation in the dorsomedial prefrontal cortex, angular gyrus, amygdala, cerebellum, and brain stem. Importantly, the present results could not be explained by prevailing levels of cardiovascular disease risk or excessive head motion, suggesting that the observed associations are not artifactual. These novel findings are consistent with the possibility that part of the relationship between stressor-evoked brain activity and preclinical atherosclerosis may be accounted for by individual differences in corresponding levels of stressor-evoked cardiovascular reactivity.

Our study builds upon a growing body of evidence for the possible brain systems and physiological pathways that may link psychological stress to preclinical atherosclerosis and CVD risk. We previously showed, for example, that brain activity patterns evoked by two different sets of unpleasant emotional stimuli were able to successfully predict individual differences of CA-IMT, explaining 1-3% of interindividual variability^49^. Important brain regions for these predictions included the insula, hypothalamus, brainstem and areas of the anterior cingulate and medial prefrontal cortices. Here, our brain activity patterns emerging from an fMRI stressor battery were able to explain a roughly similar variability of CA-IMT (∼2%) across individuals and particularly engaged similar brain areas. Thus, our results go in line with the supporting evidence of an interdependence between executive functions, contextual appraisals, and affective processes^50–53^, which here is also reflected in their similar relationship with preclinical atherosclerosis. It remains to be tested whether the integration across all these different task paradigms could boost the prediction of individual differences in cardiovascular disease risk, similar to what was found for blood pressure reactivity using the same two tasks employed here^33^. Importantly, integrating patterns of fMRI activity across different task paradigms could be an alternative to combining different neuroimaging modalities, as the latter has not been shown to improve the prediction of preclinical atherosclerosis^54^.

We also demonstrated that stressor-evoked brain activity relates to acute changes in systolic blood pressure, which was consistent across the three cardiovascular reactivity measures that we tested. Indeed, our supplementary results using L1-penalized principal component regression and involving ΔSBP as the outcome variable (min r=0.206, max r=0.256; see Suppl. Figure 7) followed those previously reported using one of the cohorts in the dataset^33^, although with a slight reduction in prediction performance (but within the same levels of confidence intervals). This is likely due to the increased sample size that tends to stabilize effect sizes to their generalizable true value^55,56^. Nevertheless, the obtained prediction rates were still small. It has been argued that one explanation for such small effect sizes is that task-fMRI measures are not reliable enough^57^. However, this argument is still in debate^58^. In fact, here we employed a sample size much larger than those reported in ^57^, and utilized task-aggregated multivariate patterns that have been proven to increase reliability to sustainable levels for the prediction of individual differences^35^. Therefore, albeit small, the observed associations of stressor-evoked brain activation patterns with preclinical atherosclerosis and cardiovascular reactivity may be likely to approximate their true value. It is also important to note that peripheral blood pressure responses are not entirely determined by brain systems for visceral control: such responses are likely to be additionally influenced by variation in autonomic outflow, peripheral autonomic receptor density and sensitivity (e.g., alpha- and beta-adrenergic receptor density and sensitivity), as well as other vascular determinants that could also account for unexplained variance in cardiovascular reactivity across individuals^59^.

Notably, amongst the three cardiovascular reactivity measures tested, only SBP_AUC_g_ appeared to mediate a meaningful association between stressor-evoked brain activity and CA-IMT. We speculate that this is due to this cardiovascular reactivity variable being the only one with a sizable association with CA-IMT, as shown by the results for the M-to-Y path in Figure 3. In our context, SBP_AUC_g_ summarizes the total SBP reactivity output under a series of events, here comprising SBP throughout MRI testing, whereas SBP_AUC_i_ is possibly more related to the sensitivity of the measure to variable changes and peak values over time^40^. Since the latter involves SBP changes with respect to the baseline in both incongruent and congruent conditions, and considering that the time separation between successive BP events throughout the task sequences is always the same, it is not surprising that it almost perfectly correlates with ΔSBP, which involves average changes with respect to the baseline only within the incongruent condition (PIP: 0.98, NOAH: 0.96, see Figure 1). It might be the case that each AUC-based cardiovascular reactivity measures corresponds to a particular feature of individual differences in preclinical atherosclerosis; e.g. overall intensity, as similarly encoded by AUC_g_ based measures, appears to mostly correlate with cross-sectional values, whereas acute reactions, as similarly encoded by AUC_i_ based measures, appear to be more important in explaining longitudinal changes over time^60^. By these and related considerations, it has been recommended to include all such metrics when analyzing data with repeated measures of stress physiology^40^. Nevertheless, in regard to the variance in CA-IMT accounted for by reactivity measures such as ΔSB (and SBP_AUC_i_ in extent), which we found to exhibit an average R^2^ = 0.009, 95% CI [0.009, 0.01] (or r=0.097, 95% CI [0.094, 0.101]), it is important to highlight that this closely compares to the meta-analytic effect size of 0.096 reported previously^14^. Accordingly, although small in magnitude, our observed effect sizes align with cumulative evidence and implicate additional biological pathways beyond cardiovascular reactivity that may link stress-related brain functionality to CVD risk, as well as underscoring the need in the field for large sample sizes and cross-validation methods.

Further, in our bootstrapping analysis applied to the scenario with SBP_AUC_g_ as the mediator, we found that this mediation effect was observed along 10 principal components computed from multivariate brain activation patterns. Strikingly, the coefficients of each of these components in the X-to-Y path, i.e., the *total effect* or *c* coefficients) did not show a statistical significance after correcting for multiple testing. Is it possible then that a mediation effect exists even in the absence of total effects? In the original casual step approach to mediation analysis, the first requirement for a possible mediation effect was that the X-to-Y path should be significant^42,61,62^. However, the necessity of this has been challenged over time^63,64^. Looking into our case, each principal component from the stressor-evoked brain activity patterns was a potential candidate for a mediating effect. We found that the estimated coefficients for these *indirect* effects (i.e. the *a·b* products) had all roughly opposite signs, which is one of the possible explanations as to how a mediation effect in the absence of an overall association may occur^65^. This might be due to the fact that principal component analysis basically finds orthogonal axes and therefore, their estimated coefficients also reflect this orthogonality in terms of exhibiting opposite signs in their association with the outcome variable, although we acknowledge that this assertion requires further exploration.

Also notable is the encoding weight pattern of the significant *indirect* effects, which pinpoints brain regions such as the ventromedial prefrontal cortex, anterior cingulate cortex, insula and amygdala. These areas are part of what has been termed a ‘visceral control network’, which is proposed to be involved in mediating psychological stress appraisals and simultaneously controlling cardiovascular physiology, with alterations in this network being linked to a higher risk of cardiovascular disease^9^. These brain systems have also been recently described as belonging to an interoceptive-allostatic brain network for regulating peripheral physiology by predictive or anticipatory processes^66,67^. Along similar lines, our results appear to provide evidence for a pathway whereby increased brain activity in systems encompassed by these networks associates with larger rises in stressor-evoked systolic blood pressure and preclinical atherosclerosis across people. Interestingly, we also observed a lateralization in the role of the amygdala, with increased activity in the left and right hemisphere to be positively and negatively associated with cardiovascular reactivity, respectively. This brain asymmetry in the activation of the amygdala has been exhaustively reported (see^68^ and references therein).

Finally, several limitations of our study should be noted. First, our cohorts (PIP and NOAH) included predominantly white individuals, relatively well-educated and free of major chronic illnesses and medication regimens that could have confounded interpretations of preclinical atherosclerosis and cardiovascular reactivity markers. As a result, whether our findings are relevant to demographically diverse individuals and clinical populations is still unclear. Yet, we have made the multivariate predictive patterns reported here publicly available, so they could be eventually tested in other populations and used as predictors in tasks involving different clinical and preclinical outcomes. Second, our results were obtained from task-based activation metrics and therefore, did not involve some of the methodological and psychometric advantages that morphological, task-free or resting-state-based measures could hold in the prediction of preclinical atherosclerosis, reactivity and other cardiovascular disease risk factors^69–72^. Third, in order to quantify mediation effects in an out-of-sample framework, we resorted to a explained variance-based measure for mediation analysis, which has its own limitations^73^. Efforts to find the optimal measure of effect size for mediation analysis continues to be an active area of research^74^.

Despite these limitations, the present findings provide novel cross-validated, predictive, and machine learning evidence for the possible mediating role of stressor-evoked cardiovascular reactivity in linking multivariate brain responses to acute psychological stressors and a vascular marker of preclinical atherosclerosis.

## Data Availability

Created data files and analysis scripts
are available at GitHub (https://github.com/CoAxLab/sbp-imt-mediation).

## ACKNOWLEDGMENTS

We thank Sara Boyko for her role in data collection.

