## Supplementary Figures and Tables for "Stressor-evoked brain activity, cardiovascular reactivity, and subclinical atherosclerosis in midlife adults"

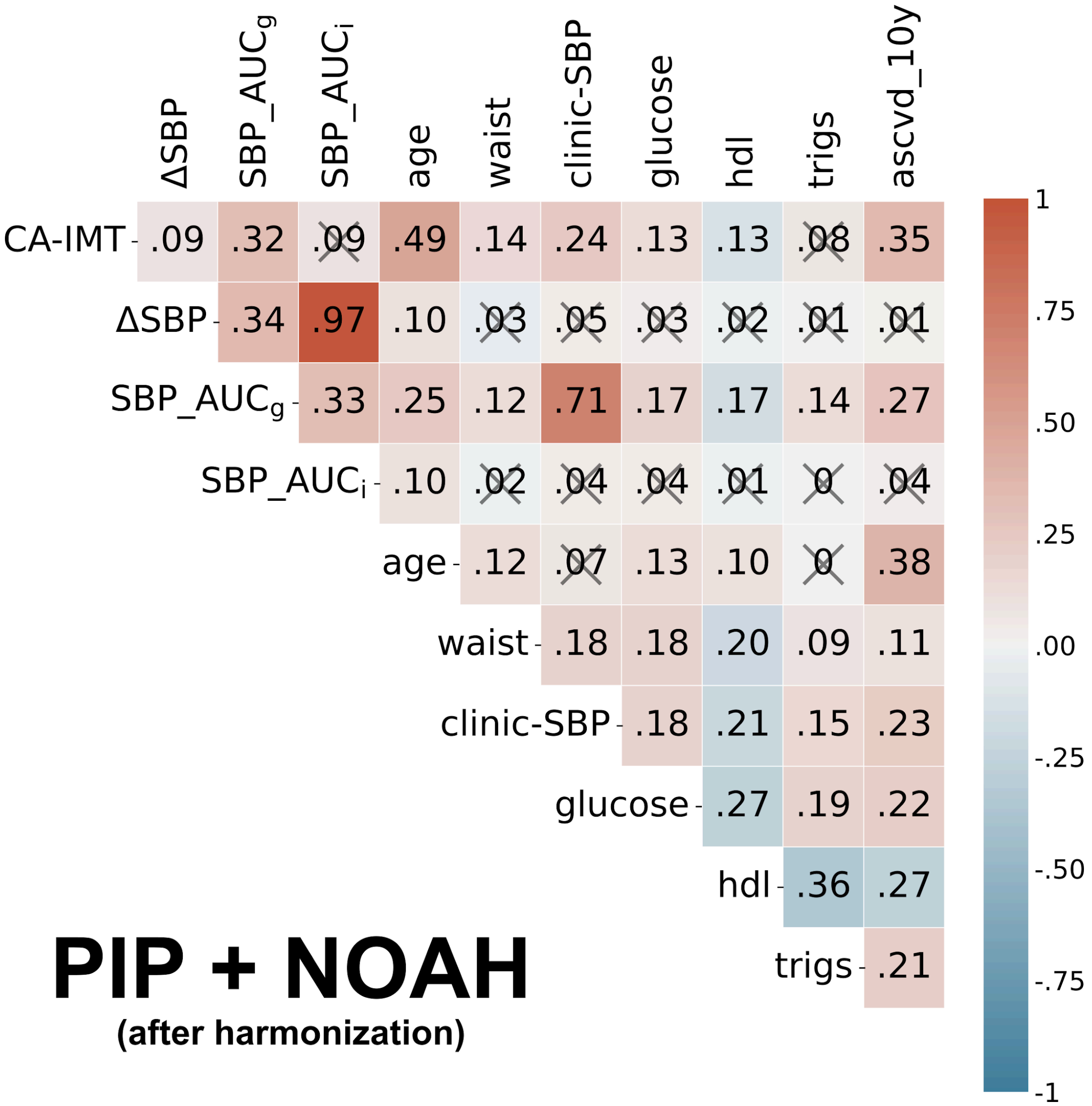


**Supplementary Figure 1.** Using the combined PIP and NOAH cohorts after harmonization, a correlogram involving the main variables in our mediation framework and several cardiovascular disease risk factors. Colors and annotations display the strength of their associations as measured by Pearson's correlation coefficients. A cross represents those associations whose p-values after false discovery rate correction were above the significance level α=0.05. SBP, systolic blood pressure; HDL, high-density lipoproteins; Trigs, triglycerides; CA-IMT, carotid artery intima-media thickness; mm, millimeter, $SBP\_AUC_g$, Area under the curve with respect to the ground; $SBP\_AUC_i$, Area under the curve with respect to increase; ascvd_10y, 10-year atherosclerosis cardiovascular disease risk score.

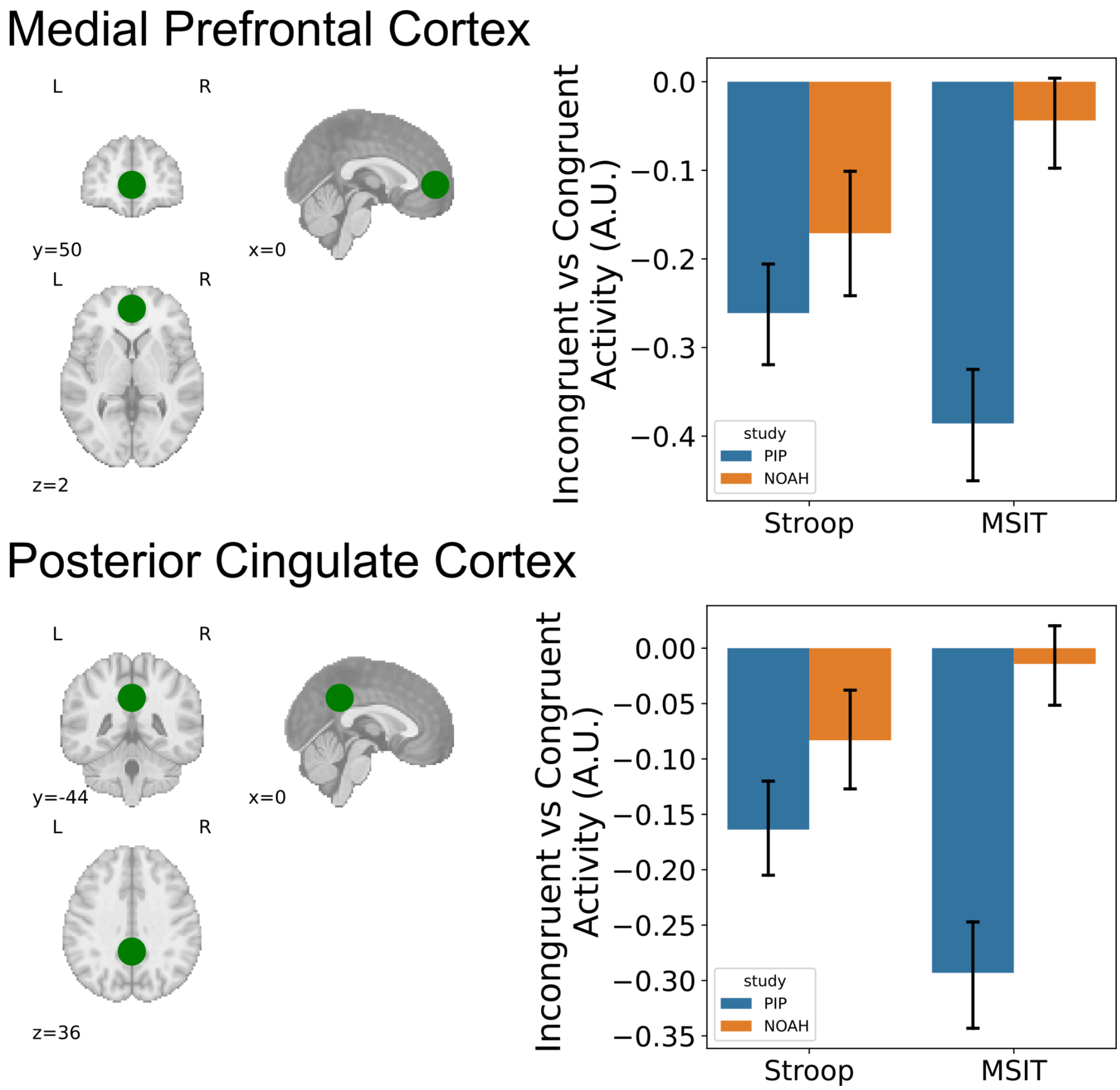


**Supplementary Figure 2.** For two different regions in the default-mode network, the brain location (left panel) and average Incongruent vs Congruent BOLD activity (right panel) in both Stroop and MSIT tasks and for both PIP and NOAH studies.

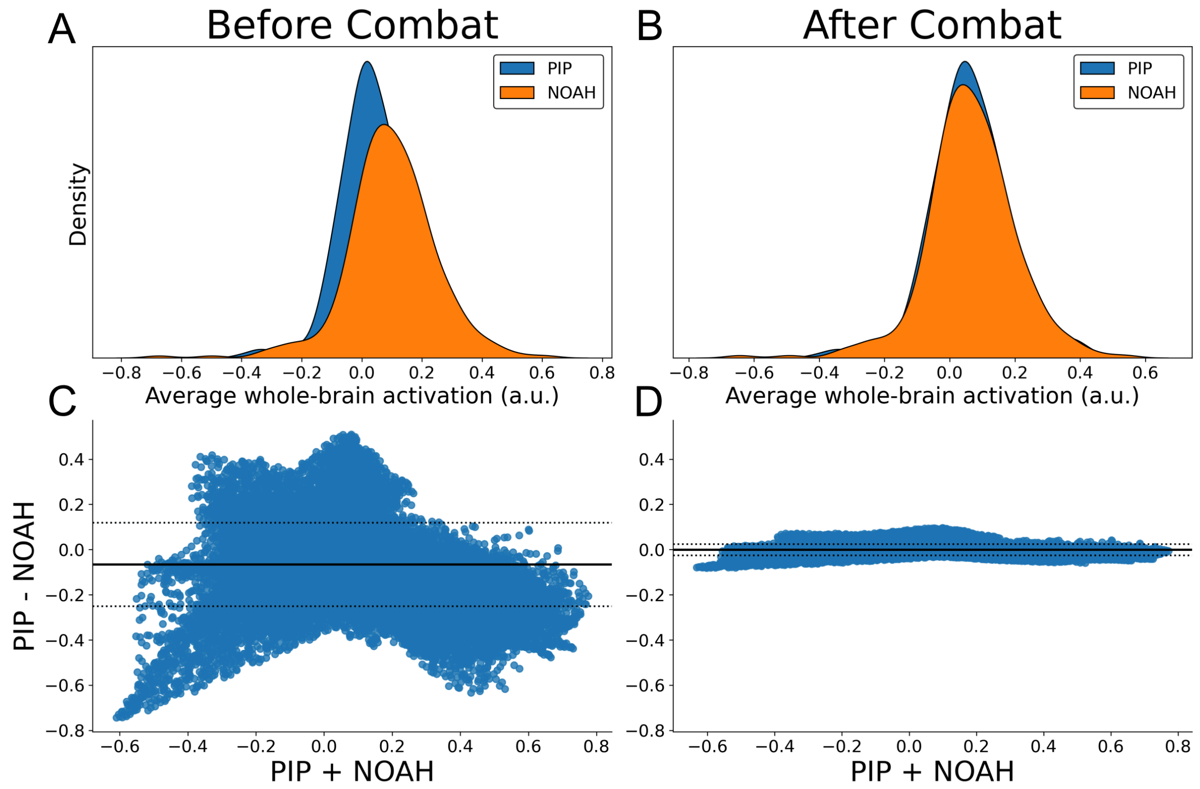


**Supplementary Figure 3.** Before (panels A and C) and after (panels B and D) Combat harmonization, the probability density plot (upper graph) using the subject-wise Incongruent vs Congruent activation, averaged over the whole brain; and the Bland-Altman plot (lower graph) for the Incongruent vs Congruent activation pattern, averaged over all subjects.

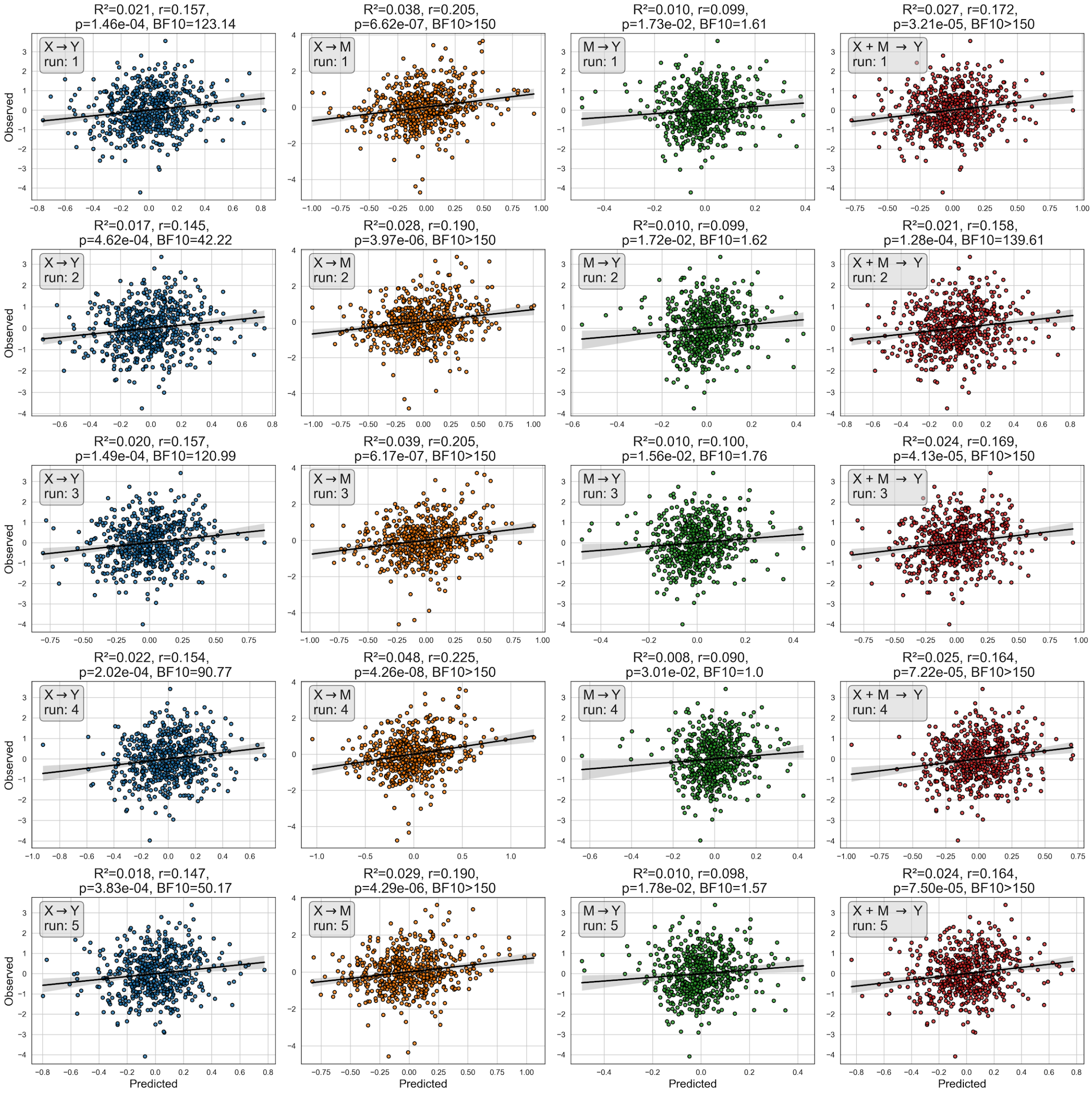


**Supplementary Figure 4.** For ΔSBP as the mediator and using a L2-penalized principal component regression, the observed .vs predicted scatter plots across the different paths in the mediation analysis framework (each column) and across the five nested cross-validation runs (each row). Dots represent the observed and predicted value for each given subject. The performance in each case is displayed at the top of the subplot, and includes the coefficient of determination $R^2$, Pearson's correlation *r*, its associated p-value, and the Bayes Factor $BF_{01}$ comparing the regression model: Observed ~ Predicted (Alternative Hypothesis) against the intercept model only: Observed ~ 1 (Null Hypothesis).

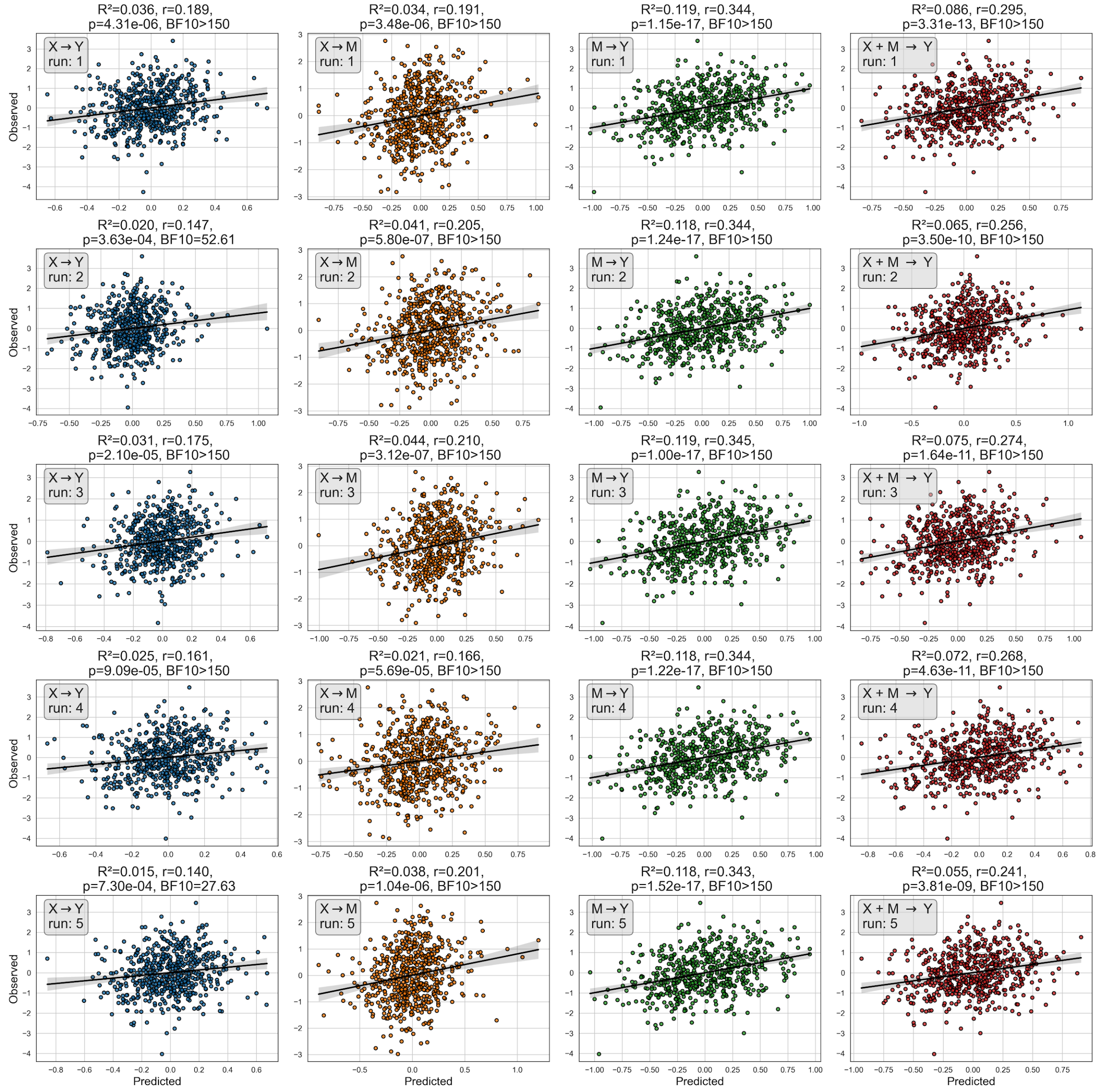


**Supplementary Figure 5.** For SBP_AUC$_g$ as the mediator and using a L2-penalized principal component regression, the observed .vs predicted scatter plots across the different paths in the mediation analysis framework (each column) and across the five nested cross-validation runs (each row). Dots represent the observed and predicted value for each given subject. The performance in each case is displayed at the top of the subplot, and includes the coefficient of determination $R^2$, Pearson's correlation *r*, its associated p-value, and the Bayes Factor $BF_{01}$ comparing the regression model: Observed ~ Predicted (Alternative Hypothesis) against the intercept model only: Observed ~ 1 (Null Hypothesis).

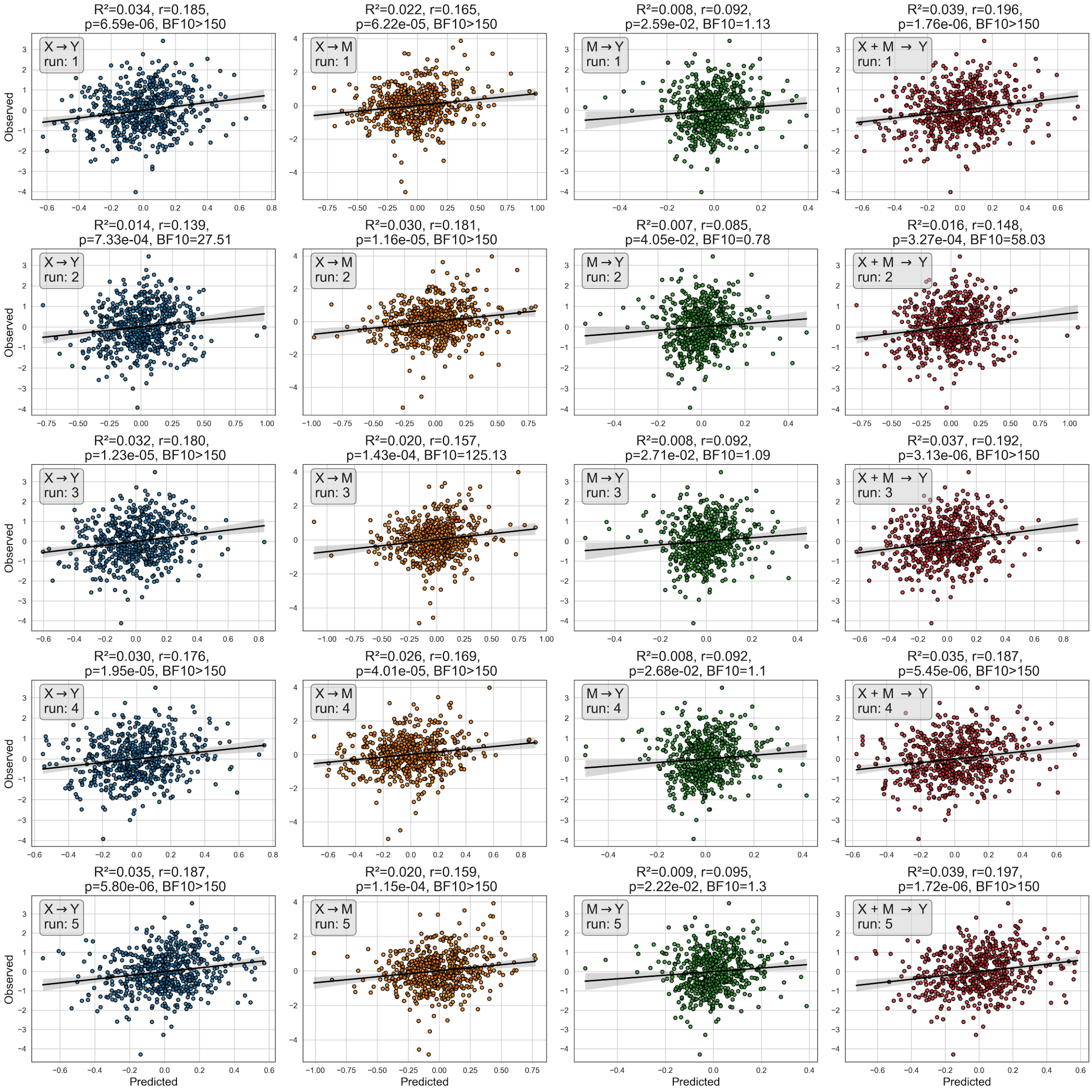


**Supplementary Figure 6.** For SBP_AUC$_i$ as the mediator and using a L2-penalized principal component regression, the observed .vs predicted scatter plots across the different paths in the mediation analysis framework (each column) and across the five nested cross-validation runs (each row). Dots represent the observed and predicted value for each given subject. The performance in each case is displayed at the top of the subplot, and includes the coefficient of determination $R^2$, Pearson's correlation *r*, its associated p-value, and the Bayes Factor $BF_{01}$ comparing the regression model: Observed ~ Predicted (Alternative Hypothesis) against the intercept model only: Observed ~ 1 (Null Hypothesis).

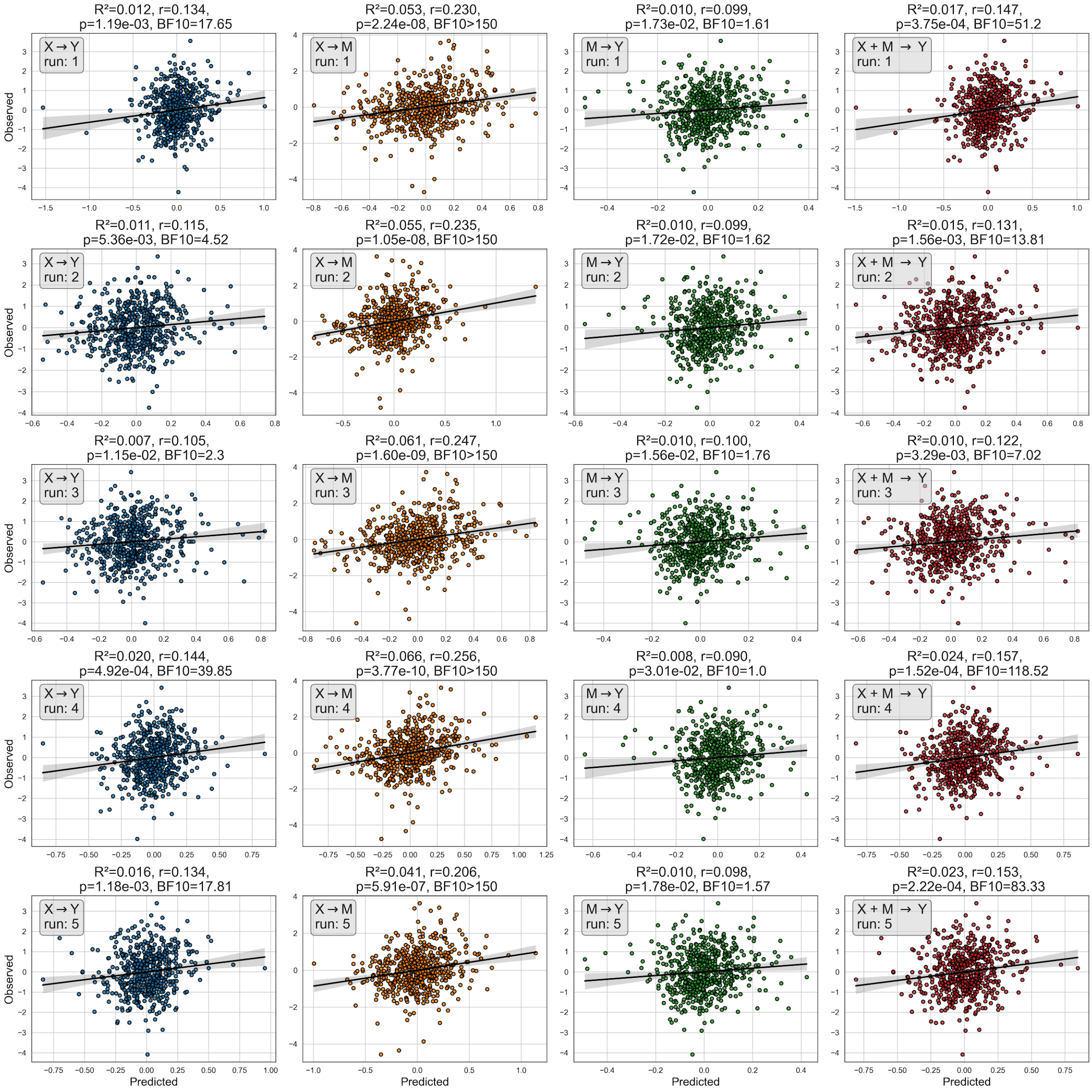


**Supplementary Figure 7.** For ΔSBP as the mediator and using a L1-penalized principal component regression, the observed .vs predicted scatter plots across the different paths in the mediation analysis framework (each column) and across the five nested cross-validation runs (each row). Dots represent the observed and predicted value for each given subject. The performance in each case is displayed at the top of the subplot, and includes the coefficient of determination $R^2$, Pearson's correlation *r*, its associated p-value, and the Bayes Factor $BF_{01}$ comparing the regression model: Observed ~ Predicted (Alternative Hypothesis) against the intercept model only: Observed ~ 1 (Null Hypothesis).

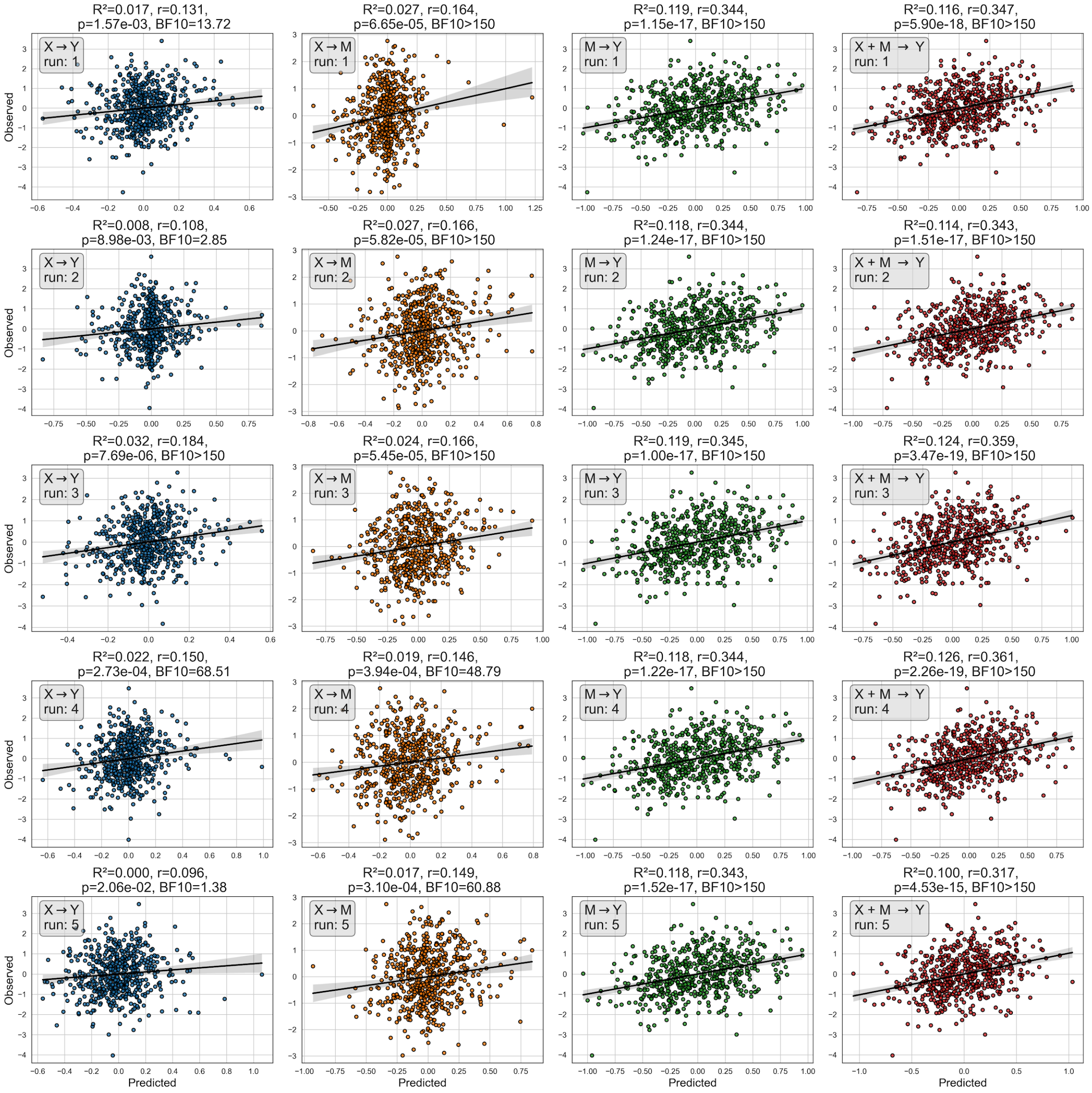


**Supplementary Figure 8.** For SBP_AUC$_g$ as the mediator and using a L1-penalized principal component regression, the observed .vs predicted scatter plots across the different paths in the mediation analysis framework (each column) and across the five nested cross-validation runs (each row). Dots represent the observed and predicted value for each given subject. The performance in each case is displayed at the top of the subplot, and includes the coefficient of determination $R^2$, Pearson's correlation *r*, its associated p-value, and the Bayes Factor $BF_{01}$ comparing the regression model: Observed ~ Predicted (Alternative Hypothesis) against the intercept model only: Observed ~ 1 (Null Hypothesis).

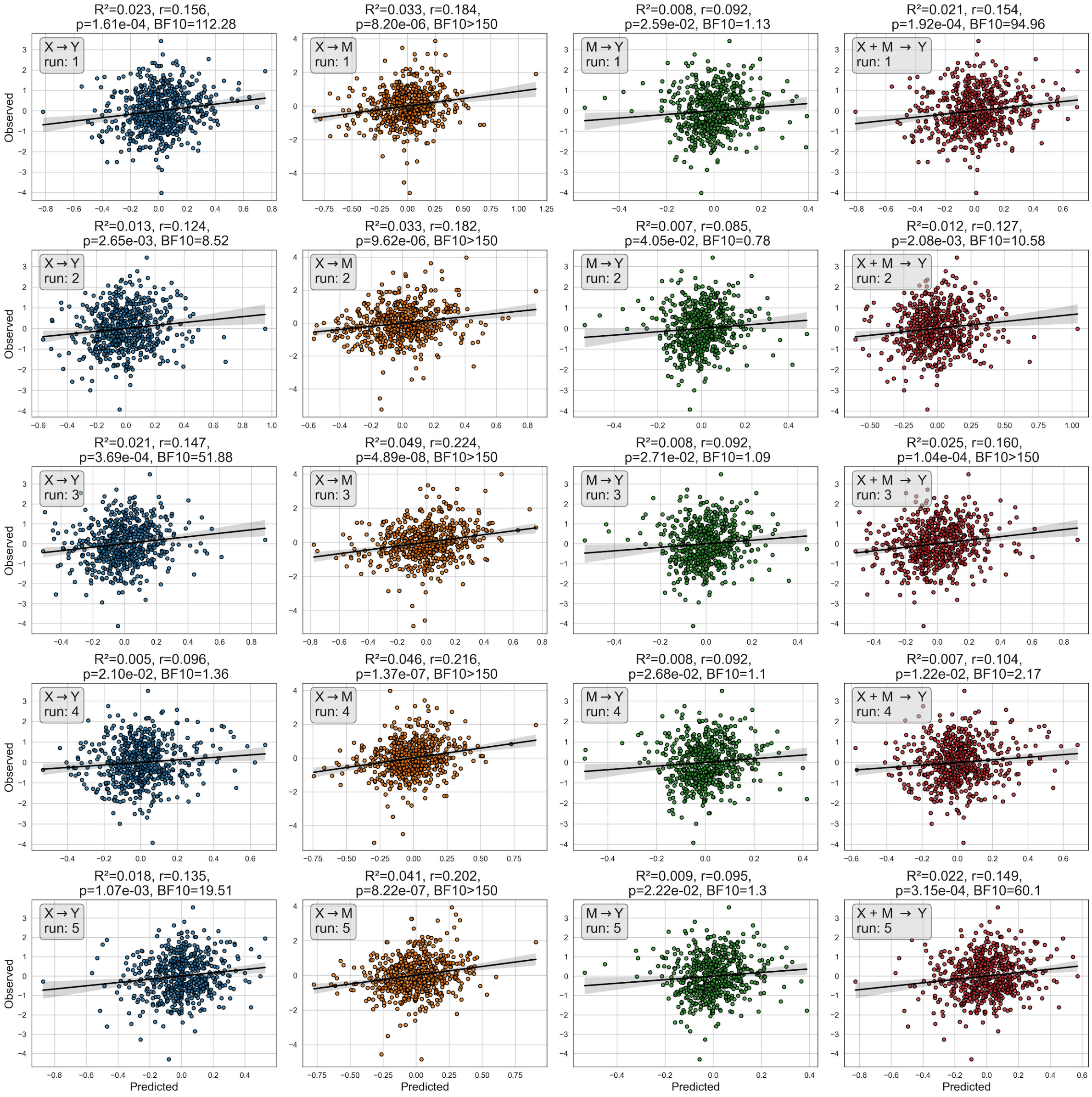


**Supplementary Figure 9.** For SBP_AUC$_i$ as the mediator and using a L1-penalized principal component regression, the observed .vs predicted scatter plots across the different paths in the mediation analysis framework (each column) and across the five nested cross-validation runs (each row). Dots represent the observed and predicted value for each given subject. The performance in each case is displayed at the top of the subplot, and includes the coefficient of determination $R^2$, Pearson's correlation *r*, its associated p-value, and the Bayes Factor $BF_{01}$ comparing the regression model: Observed ~ Predicted (Alternative Hypothesis) against the intercept model only: Observed ~ 1 (Null Hypothesis).

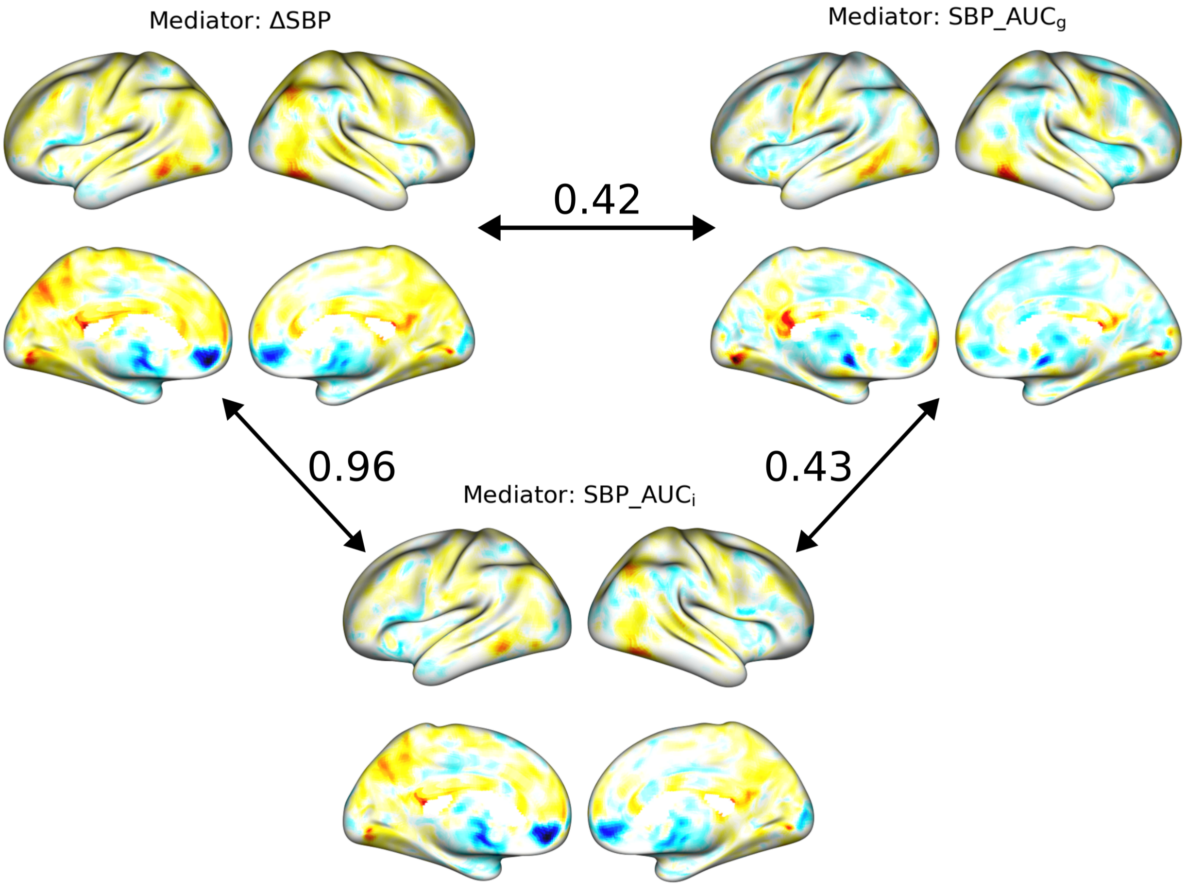


**Supplementary Figure 10. Comparison between X-to-M-based encoding weight maps.** For each mediator, the encoding weight map in the X-to-M path. In arrows the spatial similarity between these maps, measured by Pearson's correlations.

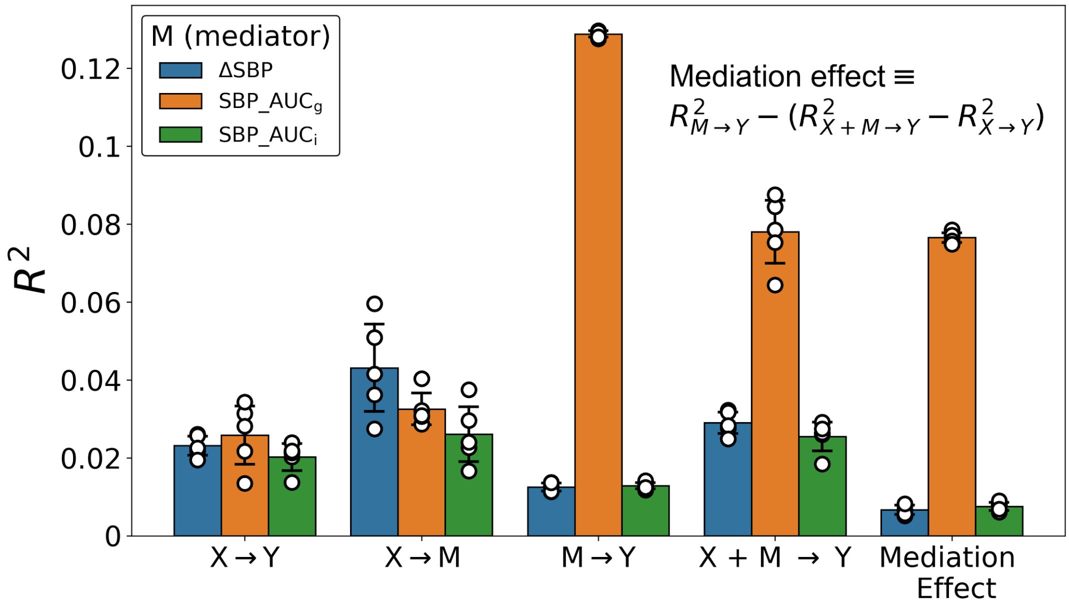


**Supplementary Figure 11. Sensitivity-analysis based-out-of-sample performances**. Concentrating exclusively on subjects with low in-scanner head motion (average framewise displacement lower than 0.5 mm in both fMRI tasks) and for each possible mediator variable, the out-of-sample performance using a L2-penalized principal component regression and applied to the different paths in the mediation analysis framework. Each dot represents the coefficient of determination calculated from the observed .vs predicted values generated from a particular run of the nested cross-validation procedure. In addition, bars and error bars display the average and the 95% Confidence intervals across these values.

| | ΔSBP | $SBP\_AUC_g$ | $SBP\_AUC_i$ |
|---|---|---|---|
| X-to-Y path | 0.152<br>[0.147- 0.157] | 0.162<br>[0.145 - 0.18] | 0.173<br>[0.156 - 0.191] |
| X-to-M path | 0.203<br>[0.19 - 0.216] | 0.195<br>[0.179 - 0.21] | 0.166<br>[0.158 - 0.174] |
| M-to-Y path | 0.097<br>[0.094 - 0.101] | 0.344<br>[0.343 - 0.345] | 0.091<br>[0.088 - 0.094] |
| [X+M]-to-Y path | 0.165<br>[0.161 - 0.17] | 0.267<br>[0.249 - 0.285] | 0.184<br>[0.166 - 0.202] |

**Supp. Table 1. Pearson's correlations.** Average Pearson's correlation and 95% CI for each mediator variable (columns) and model path (rows).
